# Identifying key multifunctional components shared by critical cancer and normal liver pathways via sparseGMM

**DOI:** 10.1101/2022.05.13.22275059

**Authors:** Shaimaa Bakr, Kevin Brennan, Pritam Mukherjee, Josepmaria Argemi, Mikel Hernaez, Olivier Gevaert

**Affiliations:** Department of Electrical Engineering, Stanford University, Stanford, CA 94305, USA; Stanford Center for Biomedical Informatics Research, Department of Medicine and Biomedical Data Science, Stanford University, Stanford, CA 94305, USA; Department of Radiology, Stanford University, Stanford, CA 94305, USA; Liver Unit, Clinica Universidad de Navarra, Hepatology Program, Center for Applied Medical Research, 31008 Pamplona, Navarra, Spain; Center for Applied Medical Research, University of Navarra, 31009 Pamplona, Navarra, Spain

## Abstract

Despite the abundance of multi-modal data, suitable statistical models that can improve our understanding of diseases with genetic underpinnings are challenging to develop. Here we present SparseGMM, a novel statistical approach for gene regulatory network discovery. SparseGMM uniquely uses latent variable modeling with sparsity constraints regulators to learn gaussian mixtures from multi-omic data. By combining co-expression patterns with a Bayesian framework, sparseGMM quantitatively measures confidence in regulators and uncertainty in target gene assignment by computing gene entropy. We apply SparseGMM to liver cancer and normal liver tissue data and evaluate the discovered gene modules in an independent scRNA-seq dataset. sparseGMM identifies PROCR as a regulator of angiogenesis, and PDCD1LG2 and HNF4A as regulators of immune response and blood coagulation in cancer, respectively. Additionally, we show that more genes have significantly higher entropy in cancer compared to normal liver; among high entropy genes are key multifunctional components shared by critical pathways, such as p53 and estrogen signaling.

**Software availability:** The software is available at https://hub.docker.com/r/shaimaabakr/sparse_gmm

**One-sentence summary:** A novel statistical approach for gene regulatory network discovery recovers modules and corresponding regulators of diverse normal liver functions, important liver cancer processes, as well as shared biology between liver cancer and normal tissue.

## 1. INTRODUCTION

Many diseases have significant genetic underpinnings that determine both the underlying pathology and potential targets for therapy. One important example where an understanding of the molecular mechanism can aid treatment is cancer. Cancer is a disease of the genome, whereby genetic and epigenetic events in certain genes, referred to as driver genes, are causal of a specific cell status that escapes normal physiological regulation and immune surveillance, leading to cancer. Altered driver genes cause dysregulation of biological pathways, downstream changes in gene expression, and cell signaling in a manner that increases cell growth and proliferation. Typically, cancer driver genes fall under the classes of master regulators, such as transcription factors, DNA-damage repair, and cell cycle genes, among others. With the high rate of genetic mutations in cancer, identifying cancer driver genes represents an important challenge. The introduction of new molecular technologies in the 2010s, such as next-generation sequencing, resulted in a surge in the availability of genomic and transcriptomic data. This increasing availability of multi-modal data is exemplified by public projects such as The Cancer Genome Atlas(*1*) (TCGA), a large-scale genome sequencing collaborative effort that aims to accelerate our understanding of the molecular basis of cancer. In TCGA, over 10,000 primary cancer and matched normal samples were characterized, spanning 33 cancer types, generating over 2.5 petabytes of genomic, epigenomic, transcriptomic, and proteomic data. Similarly, the Genotype-Tissue Expression(*2*) (GTEx) is a resource of genetic variation and expression of 54 tissue types in a large population of healthy individuals. Although the GTEx subject population does not contain disease samples, understanding the genetic and genomic variations in healthy tissue can help gain useful insight into genetic diseases and their molecular features. For instance, GTEx data was used to identify the role of a novel coronary artery disease risk gene(*3*), for detecting pathogenic gene variants related to rare genetic disorders(*4*), and GTEx data was successfully combined with TCGA data to develop prognostic markers of Acute myeloid leukemia (AML)(*5*). Leveraging these large size multi-modal data sets to make significant biological discoveries and extract clinically actionable information is only possible through developing suitable statistical models and machine learning algorithms.

Gene regulatory networks (GRNs) are one class of tools that can be applied to genomic data to improve our understanding of systems biology and uncover the molecular basis of disease. Network methods can be used to model gene-level relationships, protein-protein and cell-cell interactions. Several approaches to learning GRNs exist including graph(*6, 7*) and module-based methods (*8-10*). In graph methods, a graph is created based on the expression data and then the graph is analyzed to extract subnetworks, with hub genes assumed to be regulators of target genes in these subnetworks. Hub genes are a subset of highly connected genes, relative to the other, less connected, downstream targets. Such scale-free network structure mimics the nature of biological networks. Graph methods were used to discover major gene hubs in human B cells(*6*). They were also used to identify new molecular targets in glioblastoma(*11*). Module-based methods typically cluster co-expressed genes directly into gene modules and as a second step identify regulators of these gene modules. Example of module-based methods include CONEXIC^(*12*)^, AMARETTO(*13, 14*) and CaMoDi(*10*), which have been shown to be more robust and better recapitulate underlying biology than graph-based methods (*8*). In a previous work, we developed AMARETTO(*13, 14*), a module-based tool that clusters co-expressed genes and assigns each module to its regulators using sparse linear regression. AMARETTO outperforms other methods in its ability to leverage information from copy number variation and methylation data to improve the discovery of regulators and their assignment to gene modules. The genomic and epigenetic events inform the choice of candidate drive genes, which are used then as features selected by sparse linear regression (LASSO). The resulting modules are functionally annotated using Gene Set Enrichment Analysis (GSEA)(*15*) techniques, elucidating the role of driver genes in cancer development and progression. In later work(*9*), AMARETTO was extended to construct a pancancer module network that confirms the common cancer pathways in different cancer types and uncovers a driver gene of smoking-induced cancers, as well as another driver gene involved in anti-viral immune response exhibited by some cancers. AMARETTO has also been extended to linking genomic and imaging phenotypes from cellular and tissue images(*16*).

In this work, we present sparseGMM, a module network approach in a Bayesian framework, whereby the clustering of target genes and the assignment of regulators are combined in one step, which allows genes to be associated to multiple modules simultaneously. The assignment of a regulator to its modules can be thus calculated with a confidence interval. More specifically, we use Gaussian mixture model (GMM) inference, where the mixture mean is represented as a weighted sparse vector of regulator expression level. This novel framework tackles an important limitation in module-based methods by allowing probabilistic assignments of target genes to modules and significance estimates of individual regulator coefficients. We show an improved performance in sparsity, compared to previous methods, choosing fewer genes as true regulators, and confirming biological knowledge of the scale-free nature of gene networks.

We apply this new algorithm to GTEx data from healthy liver tissue, as well as hepatocellular carcinoma (HCC) samples from TCGA. Our algorithm can recover healthy tissue modules such as energy metabolism pathways and cancer specific modules involved in antigen presentation, immune response, and blood coagulation. We also discover common modules in healthy liver and hepatocellular carcinoma responsible for inflammation and steroid biosynthesis, among others. Further, we use a publicly available single cell data set of CD45+ immune cells(*17*) to evaluate immune related modules discovered using the bulk sequencing data. The single cell evaluation of immune modules was able to decouple distinct myeloid and lymphoid biological processes in the HCC micro-environment. Our results demonstrate the ability of our method to represent GRNs as potentially overlapping gene modules as demonstrated on bulk and single cell RNA seq data.

Further, contrary to previous methods, the probabilistic assignment approach taken by sparseGMM is potentially superior for modeling genes with multiple biological functions. Thus, we define the entropy of a gene to be the entropy of the estimated module-assignment probability, and show that it can then be used as an indicator of a multifunctional biological role based on joint membership to two or more modules. These multifunctional genes could in turn translate to multifunctional proteins having central roles in the crosstalk between two or more pathways in cancer cells, and, thus become attractive targets for overcoming drug resistance through compensation mechanisms. We show that high-entropy genes are more common in cancer samples than in healthy tissue, and we associate them to crosstalk between several pathways including TP53, interferon gamma and TNF alpha. Our analysis of high entropy genes exemplifies ways in which major cancer pathways share key multifunctional components.

## 2. RESULTS

Here, we present a new method, sparseGMM, which uses a Bayesian latent variable approach to model the relationship between regulators and downstream target genes (see Methods, Supplementary Note 2). To validate our approach, we apply our method to two bulk gene expression liver data sets of normal liver and liver. Gene modules in normal tissue were constructed using publicly available data from GTEx project, while cancer modules were constructed using hepatocellular carcinoma data from the TCGA project (Figure *1*). We used community detection methods to screen gene modules for robustness and to uncover shared biology between normal liver tissue and liver cancer. Next, we evaluated these communities in an independent single cell data set containing CD45+ immune cells from HCC cancer patients and analyzed the expression of these communities in different immune cell populations.

**Figure 1:**
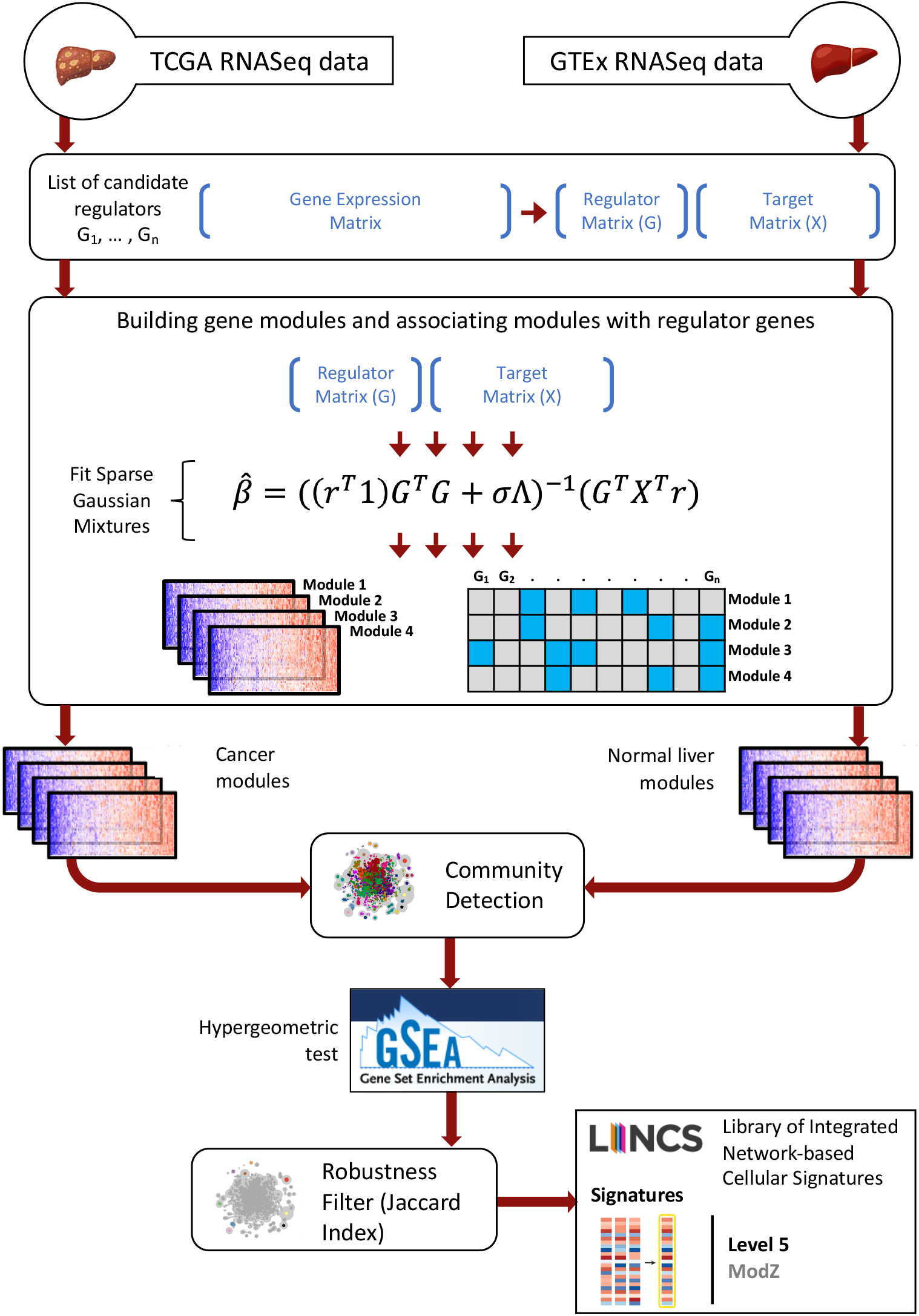
Overview of study and the SparseGMM method. SparseGMM uses a graph-based Bayesian framework combined with co-expression pattern to connect sparse sets of regulators to their downstream target gene modules. To measure robustness, we ran sparseGMM several times generating multiple gene networks from each of two data set with normal liver and liver cancer gene expression profiles. To screen for robust modules and identify normal-cancer shared biology, we ran a community detection algorithm to group robust modules that are consistently discovered in every run. Next, we performed functional gene set enrichment analysis using MSigDB gene collections. Finally, we used publicly available perturbation experiments that identify experimental targets to validate SparseGMM regulators.

### 2.1. Technical validation

For both TCGA(*1*) and GTEx(*2*) data, we compared sparseGMM to AMARETTO(*13*) and showed improved performance in terms of sparsity of regulators for various choices of the regularization parameter values (Figure *2*, Supplementary Table 1). AMARETTO was selected as, to the knowledge of the authors, it is the current state-of-the-art method for module-based GRN inference. Analyzing sparsity performance in GTEx and TCGA data, sparseGMM outperforms AMARETTO for all choices of the regularization parameter, lambda, with sparser solutions being more desirable(*18*). The mean number of regulators per module, with sparsity parameter lambda=500, is 21.27 for sparseGMM, comparted to 84.14 for AMARETTO using GTEx data (p-value<0.05, independent t-test). Similarly using TCGA data, the mean number of regulators is 38.20 for sparseGMM comparted to 196.33 for AMARETTO (p-value<0.05, independent t-test). For robustness measured using the Adjusted Rand Index (ARI) on modules from multiple runs on each data set, both data sets show similar performance with an increasing trend as the regularization parameter increases. On the other hand, both methods show gradual decrease in R^2^ with increased regularization for both data sets. SparseGMM performs better than AMARETTO for lower values of lambda. Module size increases with regularization for both data sets. At lambda=5e3, sparseGMM has larger module sizes than AMARETTO. At values of lambda>500, the module sizes are too large for practical functional annotation and discovery. Overall, the sparsity performance of sparseGMM was superior for all tested values of the regularization parameter. Acceptable module sizes and R-squared were seen for lambda=500 and lower similarly, while acceptable adjusted Rand index values were seen at 500 and higher. These results dictated the choice of lambda=500 in subsequent analyses.

**Figure 2:**
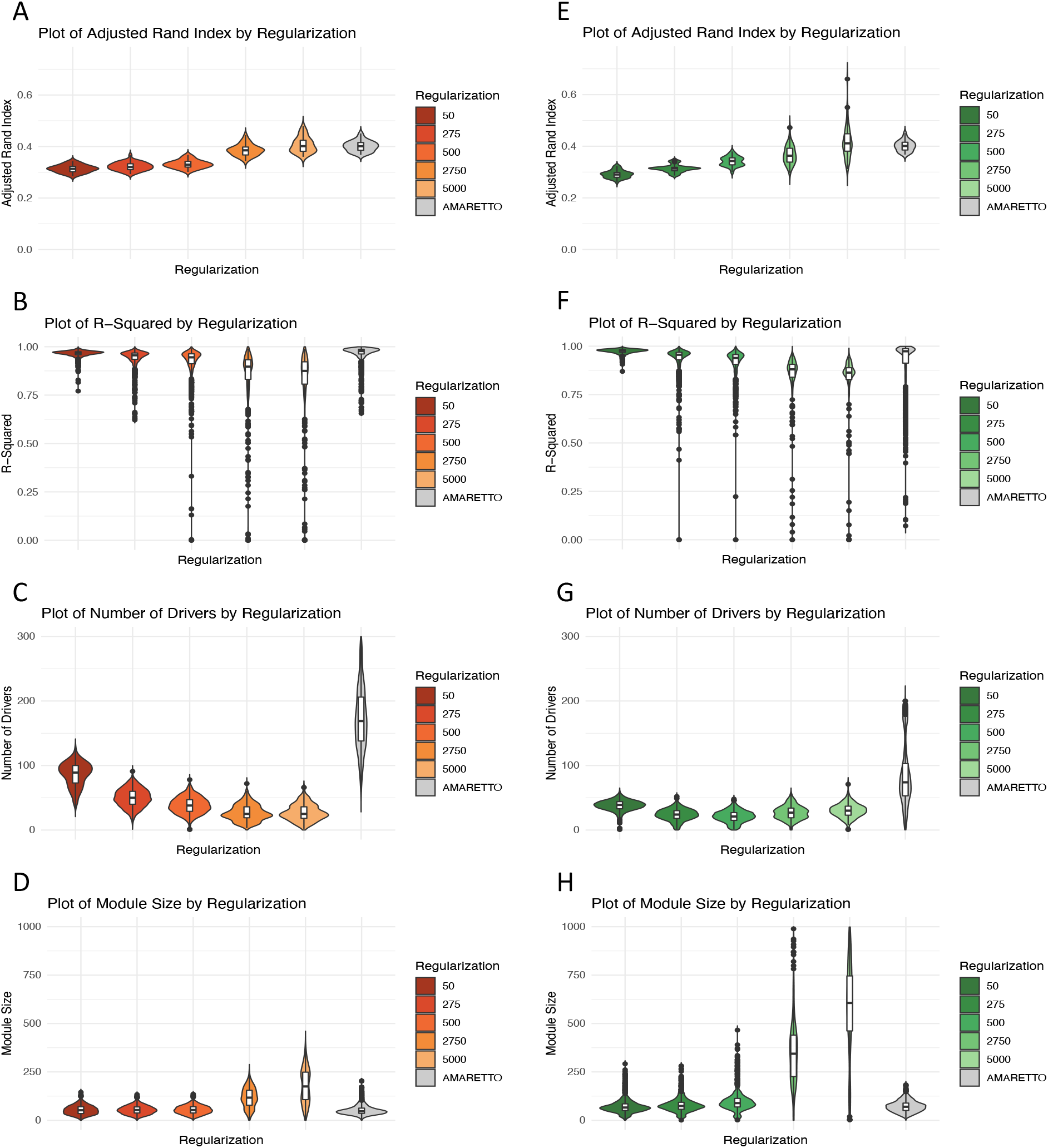
Performance comparison between SparseGMM and AMARETTO at different regularization values. Comparison shown for TCGA HCC (A-D) and GTEx (E-H) liver data. (A,E) Robustness of clustering is evaluated using adjusted Rand index. (B,F) Validation of regulators is represented by R-squared. (C,G) Degree of sparsity is evaluated using statistics on the number of drivers. (D,H) Module size informs the choice of regulatization parameter value.

### 2.2. Liver cancer and healthy livers share an angiogenesis community

From the combined analysis of normal liver and liver cancer tissue, we discovered 72 communities containing normal liver modules, cancer modules, and communities that combined normal and cancer modules (Figure 3). We defined robust communities to be those with an average pairwise Jaccard Index>=0.7 between each two modules. We found 22/72 such communities and were able to reliably identify the biological function of 15/22 robust communities (See Methods, Supplementary Table 2). We used the Library of Integrated Network-Based Cellular Signatures (LINCS) database to validate the uncovered regulatory relationships. Although many of the regulators do not have corresponding LINCS perturbation experiments, 9 communities (out of 11 non-immune highly robust communities) had at least one regulator validated using LINCS perturbation experiments. For immune communities, we were able to identify known regulators using evidence from previous studies (Supplementary Note 1). We hypothesized that sparseGMM could be useful to find communities shared by both HCC and healthy tissues, leading to the identification of highly conserved functions in HCC. We, therefore, investigated the shared GTEx and TCGA modules, revealing four robust communities enriched in functions important for physiological liver regeneration upon damage and tumor growth, including angiogenesis, cell cycle/DNA replication, ribosome and sterol biosynthesis (Supplementary Table 2).

**Figure 3:**
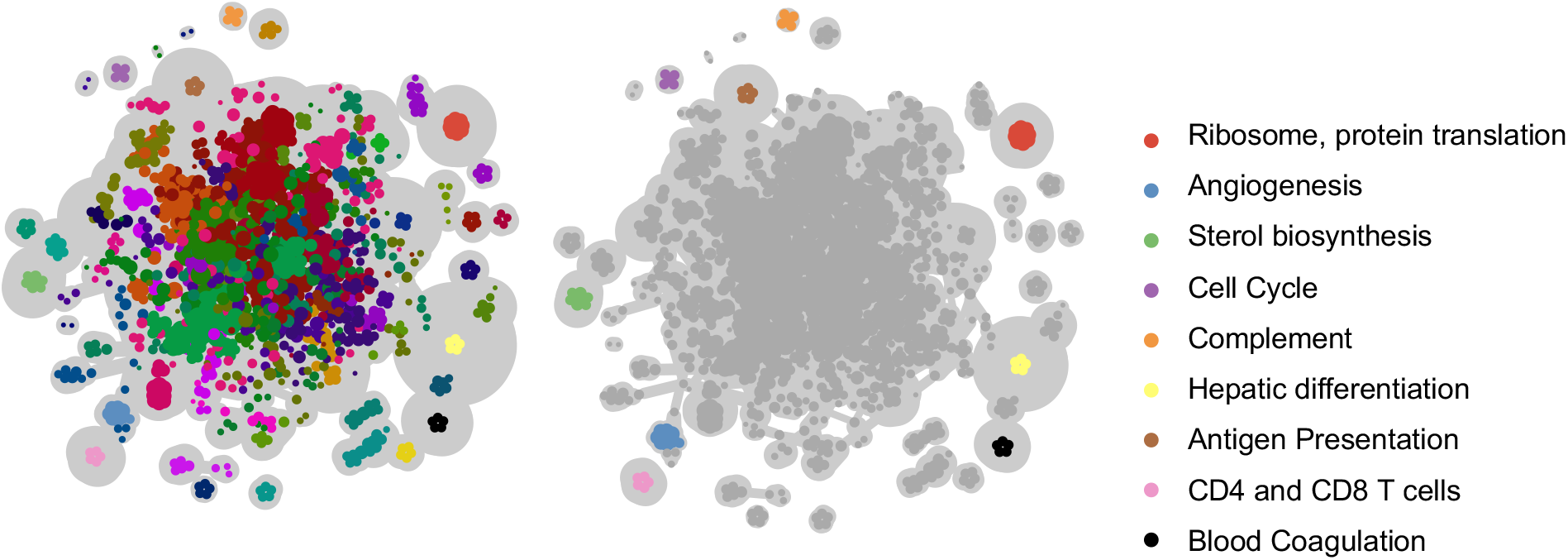
Sparse GMM module network. Left: A sample module network obtained through community detection algorithm to cancer and normal liver modules, after running sparseGMM with different initializations. Right: The community detection clusters robust modules together into distinct subnetworks. Subnetworks at the periphery represent robust modules. Subnetworks are then functionally annotated using gene set enrichments analysis applied to MSigDB gene sets. Highlighted here are robust modules from normal liver, liver cancer, as well as shared communities that contain modules occurring in normal and cancer tissue.

We highlight a shared angiogenesis community that is enriched in gene sets that relate to vasculature development, extension of new blood vessels from existing capillaries into vascular tissues and movement of an endothelial cell to form an endothelium. LINCS perturbation data confirms 2/2 regulators (Table 1). The first is NPDC1, a neural factor, which down-regulates cell proliferation(*19*). Secondly, PROCR(*20*) is a receptor of activated protein C, which has a documented role in inhibiting metastasis(*21*) and limiting cancer cell extravasation through S1PR1(*22*). Interestingly, S1PR1 is also a regulator in this community with well-documented roles in angiogenesis and liver fibrosis(*23-26*), but was not validated due to lack of perturbation experimental data in the LINCS database. PROCR was also shown to induce endothelial cell proliferation and angiogenesis(*27*) and identified as a biomarker of blood vascular endothelial stem cells(*28*) and a potential cancer biomarker(*29*). Among the shared regulators between cancer and normal samples is LDB2, a transcription factor, which regulates the expression of DLL4(*30, 31*), a notch ligand Involved in angiogenesis; DLL4 negatively regulates endothelial cell proliferation and migration and angiogenic sprouting(*31*).

**Table 1:**
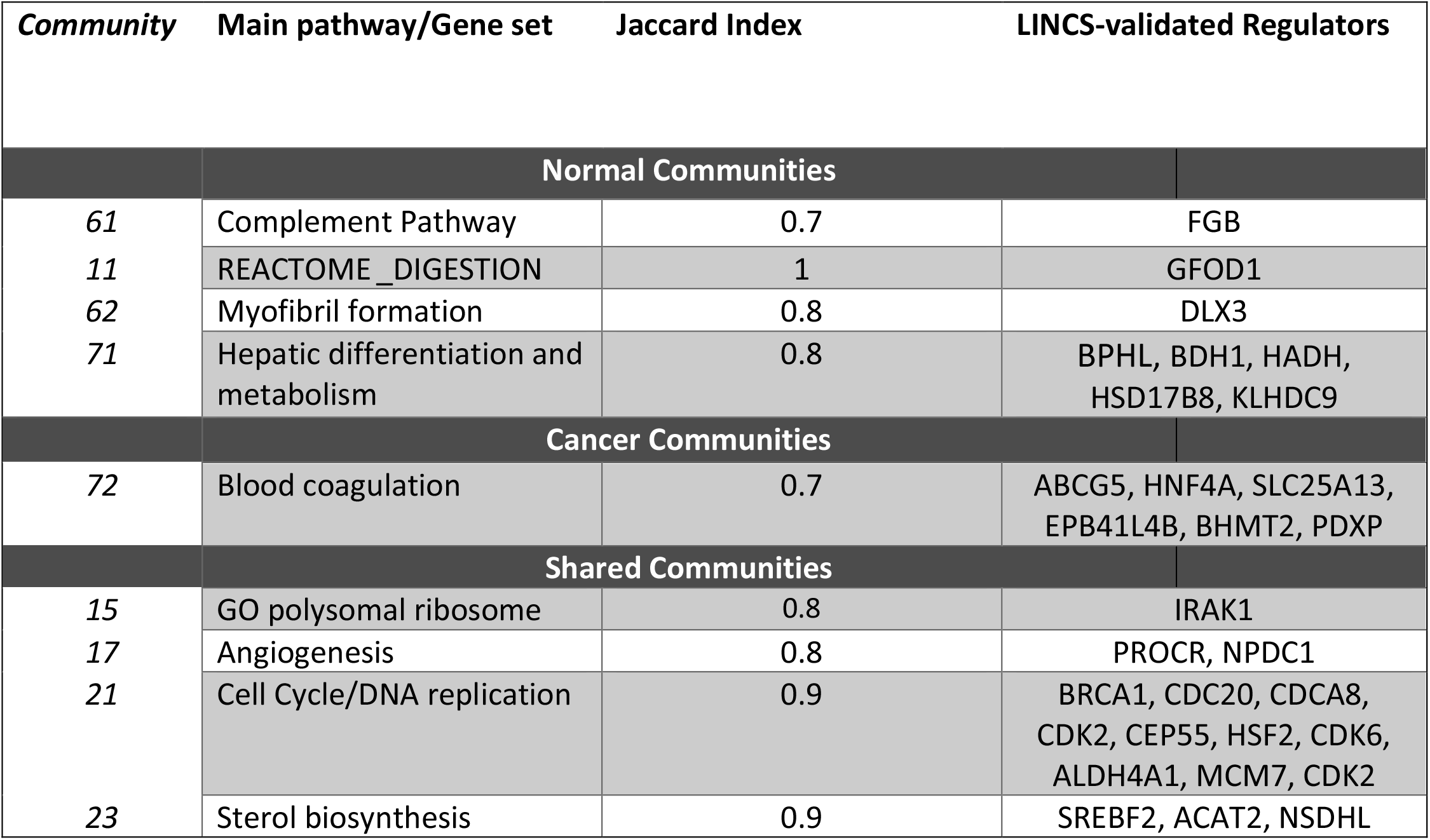
LINCS validation of robust normal liver and liver cancer communities. Robust communities were defined by having Jaccard Index >= 0.7. Main pathway of each community was revealed through gene set enrichment analysis of sparseGMM modules in GTEx and TCGA data against MSigDB collections. Validation of regulators is established with an adjusted p-value < 0.05.

| **Community** | **Main pathway/Gene set** | **Jaccard Index** | **LINCS-validated Regulators** |
| --- | --- | --- | --- |
| **Normal Communities** |  |  |  |
| 61 | Complement Pathway | 0.7 | FGB |
| 11 | REACTOME_DIGESTION | 1 | GFOD1 |
| 62 | Myofibril formation | 0.8 | DLX3 |
| 71 | Hepatic differentiation and metabolism | 0.8 | BPHL, BDH1, HADH, HSD17B8, KLHDC9 |
| **Cancer Communities** |  |  |  |
| 72 | Blood coagulation | 0.7 | ABCG5, HNF4A, SLC25A13, EPB41L4B, BHMT2, PDXP |
| **Shared Communities** |  |  |  |
| 15 | GO polysomal ribosome | 0.8 | IRAK1 |
| 17 | Angiogenesis | 0.8 | PROCR, NPDC1 |
| 21 | Cell Cycle/DNA replication | 0.9 | BRCA1, CDC20, CDCA8, CDK2, CEP55, HSF2, CDK6, ALDH4A1, MCM7, CDK2 |
| 23 | Sterol biosynthesis | 0.9 | SREBF2, ACAT2, NSDHL |

### 2.3. Antigen presentation and blood coagulation are robust communities revealed by sparseGMM in HCC

After applying SparseGMM only to HCC gene expression data, we discovered five robust communities enriched in pathways with important roles in the interaction between hepatocytes and the immune system: antigen presentation, interferon signaling, myeloid and CD4 and CD8 T cells, and blood coagulation (Figure 3, Supplementary Table 2).

The antigen presentation community included 34 target genes that are directly involved in the process of antigen processing and presentation by the HLA complex to the TCR present in the surface of immune cells (Supplementary Figure 1). This community is regulated by the PDCD1LG2 gene, encoding PD-L2, an immune checkpoint receptor of PD-1 and a recently adopted revolutionary immunotherapy drug target in HCC patients. In our analyses, PDCD1LG2 appeared as one of the regulators of the myeloid community, while PDCD1 regulated the T cell community (Supplementary Table 3, Supplementary Figure 1, Supplementary Note 1).

Next, we highlight the community enriched in pathways related to components of the blood coagulation system, and the clotting cascade (Figure 4B). This community is also enriched in processes involved in the maintenance of an internal steady state of lipid and sterol, which interact with the coagulation system(*32, 33*). Of the 31 regulators in this community, LINCS experimental data was available for 13 genes and 6 (46%) genes were validated (Table 1). Among these, HNF4A is the main transcriptional regulator in hepatocytes and regulates multiple coagulation genes(*34-38*). Other validated regulators of this community include EPB41L4B, which promotes cellular adhesion, migration and motility in vitro and is reported to play a role in wound healing^(*39*),(*40*)^. SparseGMM also correctly identified SERPINC1 as a regulator of this community. While there are no LINCS perturbation experiments for SERPINC1, the regulatory role of this member of the serpin family in blood coagulation cascade has been well-documented in previous studies(*41, 42*). These results show that the clotting system is robustly regulated in HCC. While impact of impaired liver function on blood coagulation is evident, the specific role of this pathway in HCC progression is largely unexplored.

**Figure 4:**
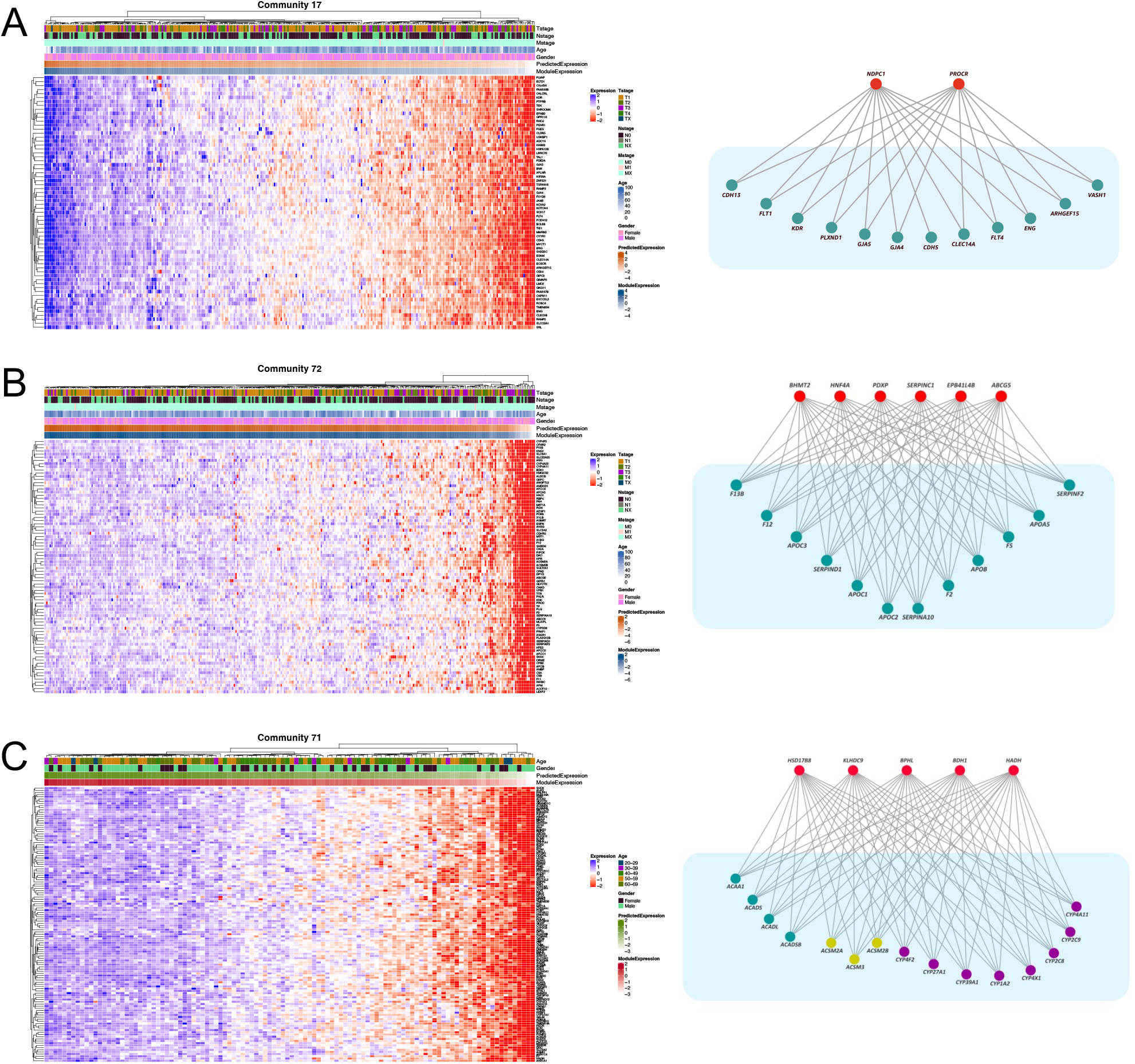
Heatmap of coexpression patters in target genes of sample modules and graph of regulatory relationships. Regulator genes are shown in red and target genes are shown in green. (A) Shared communtiy between HCC and normal liver: PROCR and NPDC1 regulate target genes of the aniogenesis community (B) Liver cancer: HNF4A and other regulators control coagulation factors and apolipoproteins involved in blood coagulation community. (C) Normal liver community: BDH1 and HADH regulate a group of Acyl-CoA Dehydrogenases and a group of cytochrome P450 enzymes involved in hepatic differentiation and metabolism.

### 2.4. SparseGMM identifies potential modules of hepatic differentiation and metabolism in healthy livers

We found six communities that highlight important normal liver functions. GSEA results reveal six distinct functions: Hepatic differentiation and metabolism, lipid and protein catabolism, complement, cancer and vesicle trafficking, myofibril formation, and FGFR1 signaling. For example, we highlight the hepatic differentiation and metabolism community, an important pathway capturing the liver’s unique metabolic functions. Specifically, LINCS perturbation experiments validated 50% of regulators (5 out of 10 with available LINCS data) in this community (Table 1). Confirmed regulators in this community include two enzymes: BDH1 a short-chain dehydrogenase that catalyzes the interconversion of ketone bodies produced during fatty acid catabolism(*43*), and HADH responsible for the oxidation of straight-chain 3-hydroxyacyl-CoAs as part of the beta-oxidation pathway(*44-46*) (Figure 4C). These findings point to sparseGMM-identified hepatic differentiation and metabolism genes as potential bona fide transcriptional biomarkers of hepatic differentiation and metabolism in healthy livers.

### 2.5. SparseGMM decouples distinct myeloid and lymphoid biological processes in HCC micro-environment, blood, and normal liver

We next evaluated the highly robust communities in an independent singe cell RNA data set of CD45+ immune cells for HCC patients from five immune-relevant sites: tumor, adjacent liver, hepatic lymph node (LN), blood, and ascites(*17*). We used Seurat to cluster the cells and compared markers of Seurat clusters to markers of various immune cells to identify the different cell types in the tumor samples of three patients (Supplementary Figure 2, see Methods). Overall, we found that 4 out of 9 communities expressed in the single cell data set were cell type specific (Figure 5A, B, Supplementary Figure 2). These communities were CD4 and CD8 T cells community, myeloid community, cell cycle community (specific to T cells and dendritic cells) and the community 60 (specific to T cells).

**Figure 5:**
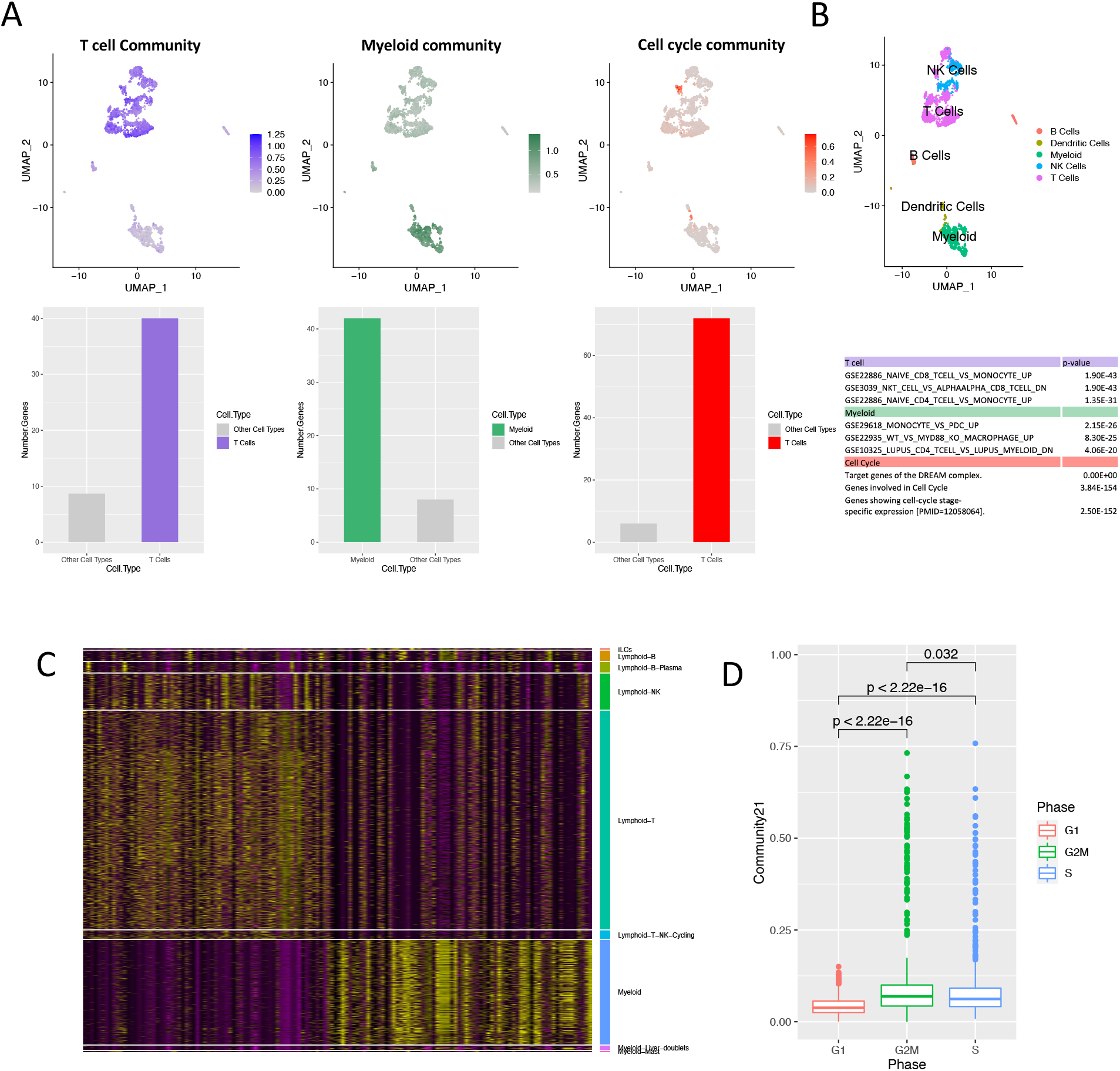
Single cell evaluation of highly robust communities. (A) Top, left to right: Average expression of the T cell, myeloid, cell cycle community and cell types. Bottom, left to right: number of genes expressed in T cell, myeloid and cell cycle community in their corresponding cell type versus average number of genes expressed in other cell types. (B)Top: Cell type annotation. Bottom: Most significant gene set enrichments for the three communities. (C) Heatmap of target genes of T cell and myeloid communities in different single cell populations. (D) Boxplot of cell cycle phase versus expression of cell cycle community target genes. Higher expression of cell cycle genes corresponds to proliferative G2M and S phases.

The expression of target genes from communities 67 and 68 distinguished CD4 and CD8 T cells from myeloid cells respectively (Figure 5A-C) with a similar expression pattern in immune cells from blood and normal liver tissue (Supplementary Figure 3). CD4 and CD8 T cells (myeloid cells) expressed a significantly larger number of genes from the CD4 and CD8 T cells community (myeloid cell community) than other cell types (adjusted p-value<0.05, chi-squared test), confirming that the communities are cell-type specific. Additionally, we observed a subset of T cells that specifically express genes from the cell cycle community (adjusted p-value<0.05 chi-squared test). As expected, the cell cycle community gene expression was lower (p-value<2.22e-16, independent t-test) in G1 phase than in the proliferating G2M and S phases (Figure 5D). When comparing this community’s average expression in cells from different environments, we found higher level of cell cycle gene expression in tumor-derived immune cells than in either normal or blood (p-value<2.22e-16, independent t-test, Supplementary Figure 3).

Finally, the percentage of variance explained in average target gene expression by regulator expression (R^2^) was 0.53, 0.80, 0.80 in CD4 and CD8 T cells, myeloid, and cell cycle communities respectively, demonstrating the accuracy of the inferred regulatory programs. These results further support the robustness of communities identified in bulk RNA-sequencing data.

### 2.6. Gene entropy identifies key elements of cancer pathway crosstalk

Both in liver physiology and liver cancer, functional crosstalk, defined as the interaction between two pathways belonging to different cell processes, are a natural way of responding to new environmental challenges. Previous studies reported crosstalk between major cancer pathways such as p53 and NF-κB/TNF-α (*47*),(*48*), and p53 and estrogen(*49*). Furthermore, this crosstalk between pathways represents compensation mechanisms by which a cancer cell can generate resistance to the blockage of a specific gene or pathway(*50, 51*). We hypothesized that gene entropy, which is a measure of uncertainty in its assignment to a gene module, could be interpreted as a proxy for multiple module membership, and thus be used to unveil the elements of hidden crosstalk in cancer.

We calculated the average entropy of each target gene over multiple runs of sparseGMM on TCGA and GTEx samples from the genes’ posterior probability (see Methods). We set an entropy threshold of 1, which corresponds to the maximum possible value of entropy between two modules, to identify genes with uncertainty in module assignment. We found that for target genes with entropy>1, TCGA target genes showed significantly higher degree of entropy when compared to GTEx (p-value<2.22e-16, independent t-test, Figure 6A). This difference in entropy distribution reflects the heterogeneity of cancer tissue compared to normal healthy tissue.

**Figure 6:**
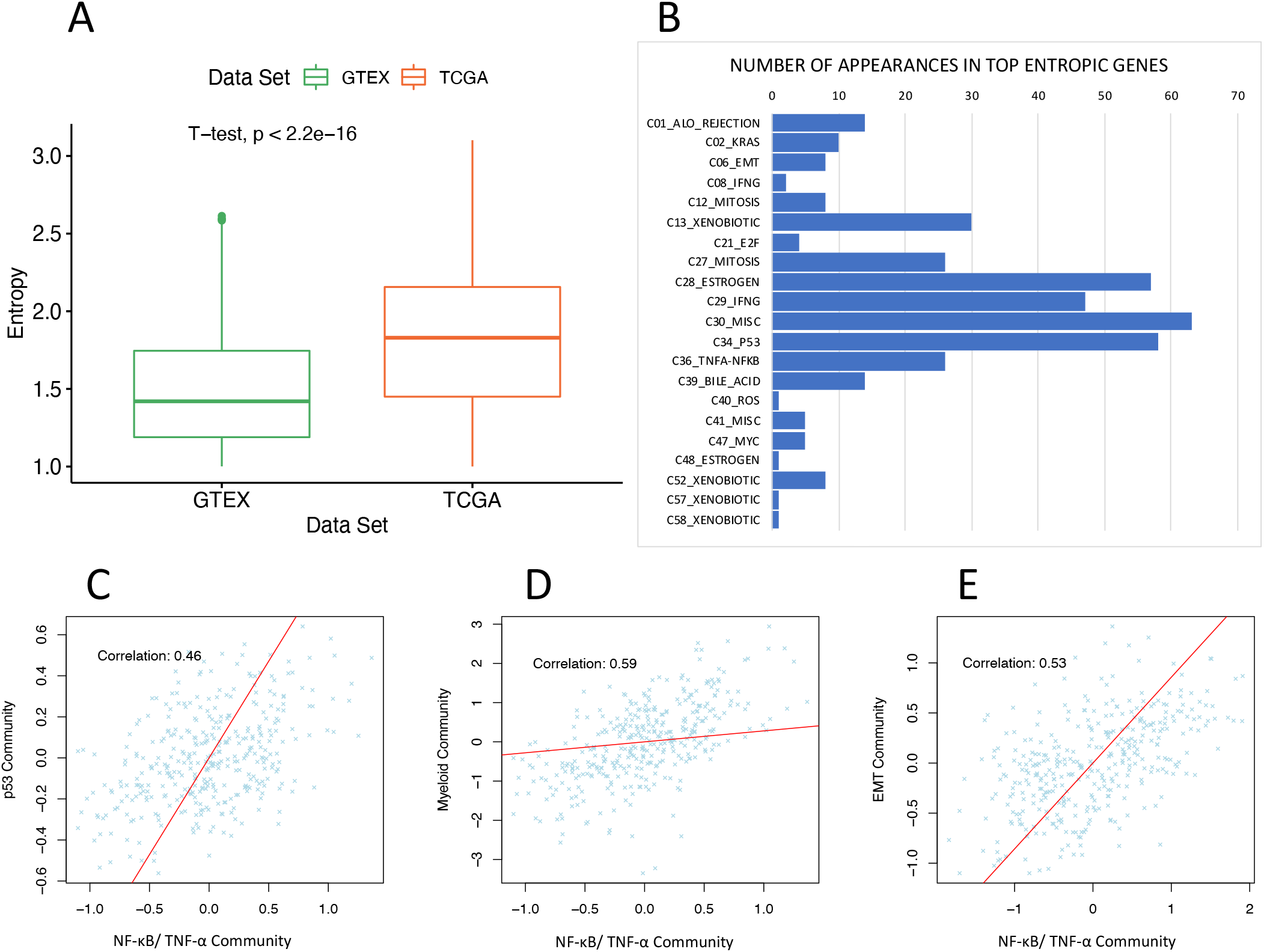
Analysis of high entropy genes. (A) Boxplot showing difference in mean entropy distribution for high entropy target genes in GTEX and TCGA, reflecting heterogeneity of cancer samples. Entropy is calculated from the posterior probability of target genes in each data set and the mean is calculated over several runs of SparseGMM on each data set. (B) Distribution of communities of high entropy genes. (C-E) Expression of communities with high entropy genes.

We then analyzed he distribution of community membership among high entropy genes in TCGA. Interestingly, genes with high entropy clustered in a few communities such as p53-related networks, NF-κB/TNF-α response, response to interferon-γ, estrogen response, and bile acid metabolism (Figure 6B). P53 harbors loss of function mutation in around one third of patients with HCC(*52*). Most HCC originate in an inflammatory liver background such as Hepatitis C or B chronic infection or Nonalcoholic steatohepatitis (NASH)(*53*), and bile acid composition has been related to HCC(*54*). Finally, estrogen signaling has been studied in liver cancer as a potential protective factor and one of the reasons HCC is more frequently seen in males than in females(*55*). Altogether, these results suggest an unbiased efficient capturing of clinically relevant pathway crosstalk by sparseGMM. If multifunctional, the genes captured by our method in each crosstalk could be important for identifying key targets for an efficient therapeutic disruption of cancer growth.

Next, we studied in detail the detected highly entropic genes within crosstalk. We found 15 high entropy genes that were assigned to both estrogen-mediated signaling and p53 communities. One of these genes, GREB1 is an estrogen-regulated gene that is expressed in estrogen receptor α (ERα)-positive breast cancer cells modulating its function and promoting cancer cell proliferation(*56*). The plausibility of a GREB1-P53 interaction is further supported by the fact that the phosphorylation of PBX Homeobox Interacting Protein 1 (HPIP), a target of p53, is necessary for estrogen-mediated GREB1 expression(*57*). Similarly, IGFALS, another high entropy gene with assignment to both p53 and estrogen signaling communities, is capable of both forming a complex with a well-known p53 target, IGFBP-3(*58, 59*), with growth inhibitory and pro-apoptotic properties(*60*), and regulating estrogen receptor in breast cancer(*61-64*).

Next, we examined more closely the crosstalk between p53 and NF-κB/ TNF-α pathways. Previous studies have shown that TNF-induced NF-κB-directed gene expression relies on p53(*48*), but the significance and specific mechanisms of this interaction are not fully explained. PAX8, a transcription factor expressed in 90% of high degree serous carcinoma(*65*), is among the highly entropic genes identified by sparseGMM as participating in both p53 and TNFa/NFkB1-related signaling. Previous studies report that while PAX8 binding is inhibited by TNF-α(*66*), its pro-proliferative role relies on p53-p21(*67*). Interestingly, we found a significant correlation between average target gene expression of p53 and NF-κB/TNF-α pathways (Figure 6C, Pearson correlation=0.46 CI [0.37-0.53], p-value < 2.2e-16). Previous studies also showed that p53 and NF-κB/ TNF-α coregulate proinflammatory gene responses in human macrophages(*68*). We observed significant correlation between the TNF-α induced NF-κB community and the myeloid community (Figure 6D, Pearson correlation=0.48, CI [0.41-0.56], p-value < 2.2e-16). The p53-NF-κB/ TNF-α crosstalk is also implicated in increased invasiveness(*69*). We found a significant correlation between the NF-κB/ TNF-α community and the Epithelial to Mesenchymal Transition (EMT) and cancer stemness community (Figure 6E, Pearson correlation=0.53, CI [0.45-0.60], p-value < 2.2e-16). Accordingly, sparseGMM is able not only to infer key regulators and communities around them, but also identify key multifunctional components shared by critical cancer pathways based on their entropy.

## 3. DISCUSSION

SparseGMM has a unique capability to infer gene regulatory network relationships from bulk RNASeq data, while assigning target genes to multiple gene modules and sparse sets of regulators to their respective modules. As a result, SparseGMM accurately models the scale-free nature of biological networks, as well as the molecular heterogeneity of biological tissue and the versatile roles of a subset of genes in different biological pathways.

SparseGMM accomplishes this goal by combining co-expression and graph-based approaches in a Bayesian setting to model the relationships between downstream target genes and their regulator genes. SparseGMM enforces a sparsity constraint, which reduces overfitting and increases the interpretability of regulatory relationships by restricting the number of regulators in each module. This sparsity in regulator selection achieves an improvement in prioritizing potential therapeutic targets and discovering new regulator genes.

We demonstrated the utility and reliability of SparseGMM by applying it to data sets of normal and cancerous liver tissue and employing community detection methods to screen gene modules for robustness and for shared biology between normal liver tissue and liver cancer. We then used gene set enrichment analysis to functionally annotate highly robust modules. Despite the complex physiology of the liver, from metabolism and immunity to protein synthesis, SparseGMM recovered important physiological functions that are active in healthy and cancerous liver tissue. SparseGMM also recovered the molecular similarities and differences between the biological processes in healthy and cancer tissues. This has implications on our understanding of the mechanisms of cancer development and progression. Further, SparseGMM was able to identify new and known regulators of normal liver physiology and hepatocarcinogenesis such as BDH1 and HNF4A. In addition, the discovery of several communities with immune function and their subsequent verification in single cell data, reflects the ability of SparseGMM to decouple biological processes related to distinct immune cell populations.

Thanks to the Bayesian nature of the proposed algorithm, SparseGMM can probabilistically assign each target gene to multiple modules and the uncertainty in gene assignment can be measured using the information theoretic measure of entropy, as a proxy for a gene’s versatile functions and capturing potential crosstalk between biological pathways. Importantly, SparseGMM identified IGFALS as a high entropy gene that was assigned to both p53 and estrogen signaling communities. In previous studies, IGFALS was reported to form a complex with IGFBP-3 a well-known target of p53(*58, 59*), which has growth inhibitory and pro-apoptotic properties(*60*). IGFALS was also reported to regulate estrogen receptor in breast cancer(*61-64*). SparseGMM was able to identify PROCR as a regulator of an angiogenesis community shared between normal liver and liver cancer. In liver cancer tissue, sparseGMM recovered an antigen processing and presentation community, regulated by PDCD1LG2, which encodes a key immunotherapy drug target in HCC and an immune checkpoint receptor of PD-1. Interestingly, PDCD1LG2 was also a regulator of the myeloid community identified by SparseGMM. In normal liver SparseGMM identified a hepatic differentiation and metabolism community regulated by BDH1, a short-chain dehydrogenase, which catalyzes the interconversion of ketone bodies produced during fatty acid catabolism(*43*). Lastly, the discovery of several communities with immune function and their subsequent verification in single cell data, reflects the ability of SparseGMM to decouple biological processes related to distinct immune cell populations. While we were able to leverage single cell data to verify immune communities, the validation of regulatory relationships in these communities cannot be performed using cancer cell line data such as LINCS data sets. Future directions include biological validation of regulatory relationships in immune communities.

In summary, SparseGMM employs a co-expression based GRN inference approach in a Bayesian framework from bulk transcriptomic data and achieves superior performance compared to state-of-the-art module-based GRN inference methods and identifies important biological pathways, and corresponding gene regulators, as exemplified by application to human liver healthy and diseased tissue.

## 4. MATERIALS AND METHODS

### 4.1. Data preprocessing

Gene modules in normal tissue were constructed using publicly available data from GTEx project, while cancer modules were constructed using HCC data from TCGA project. The data sets were preprocessed and provided in (*70*), in which they provided reference RNA-seq expression levels from healthy human tissue that can be compared with the expression levels found in human cancer tissue.

A list of candidate genes was obtained from previously generated AMARETTO(*13*) data objects extracted using the TFutils R package(*71*). In addition, genes whose gene expression can be explained using changes in copy number variation(*72*) or methylation(*73*) status from the TCGA data set were extracted using the AMARETTO package. The combined list of genes was used as an initial candidate regulator gene list. Next, the top 75% varying genes were identified to each data set separately. Of the top 75%, the top 2000 genes that are also present in the candidate regulator genes list were used to build the regulator gene matrix, the rest were regarded as target genes. The gene expression data matrix was centered to mean 0 and standard deviation 1 and then split into a regulator gene matrix and a target gene matrix. A similar approach was used to preprocess and build the input data matrices from the GTEx data set. Overall, the TCGA contained 8017 protein-coding genes including 2000 candidate drivers, while the GTEx network contains 10804 protein-coding genes including 1800 candidate drivers.

### 4.2. Implementation and technical validation

We implemented SparseGMM (Supplementary Note 2) in Python. SparseGMM was run 5 times on each data set with different seeds to evaluate the robustness of the method. We first split both data sets into training (70%) and test (30%) sets. Four different metrics were employed to validate the performance of sparseGMM: 1) Adjusted index (ARI) to measure robustness, 2) R-Squared to measure goodness of fit, 3) Number of selected regulators to measure sparsity, and 4) Size of module to evaluate the sensitivity of module size to the regularization parameter lambda. The values of lambda above 5000 produced very large modules and were excluded from further analysis. Values below 50 were also excluded due to low adjusted rand index. Lambda values examined were 50, 275, 500, 2750 and 5000 were used. The output of different seeds was also used to filter the generated modules using cAMARETTO as explained below. The input number of clusters used was 150 as it resulted an average size of 60 genes per cluster and reduced false positive results in downstream functional gene set enrichments (Figure *2*D, H).

### 4.3. Robust module recovery via community detection

After running SparseGMM we used the community AMARETTO package(*16*) to build communities among modules discovered by running sparseGMM with different seeds on the same data set. Modules that are consistently discovered by sparseGMM clustered together in the same community. cAMARETTO parameters used were p-value = 0.01 and intersection = 10. When running cAMARETTO on a single data set (either GTEx or TCGA), we filtered for communities of size 5, one from each run and further narrowed down results by Jaccard index >=0.7. In contrast, for communities with both TCGA and GTEx modules, communities of size 10 were selected. The selected communities were used as input to the GSEA function of cAMARETTO.

### 4.4. Gene set enrichment analysis

We then applied GSEA using MSigDB collections (Hallmarks and C1-5) to functionally annotate each of the communities. A p-value<1e-5, adjusted for testing of multiple hypotheses using the Benjamini-Hochberg method, was selected to filter enriched data sets.

### 4.5. Biological validation

To experimentally validate regulators of the discovered communities, we interrogated the robust regulators, defined as regulators consistently associated with the same community by sparseGMM across all runs, against publicly available genetic perturbation studies in the Library of Integrated Network-Based Cellular Signatures (LINCS) database. In this validation experiment we leveraged the HEPG2 liver cell line data. The types of perturbation experiments used were 1) Consensus signature from shRNAs targeting the same gene and 2) cDNA for overexpression of wild-type gene. The Fast Gene Set Enrichment Analysis tool was used to test for significance in enrichment. To empirically derive p-values, we permuted 1000 lists of genes of the same size as the community target sets for each community and for each regulator. Regulator-gene set pairs which had a corresponding p-value<0.05, adjusted for testing of multiple hypotheses using the Benjamini-Hochberg method, were considered validated cellular signatures in either of the two signature types.

### 4.6. Single cell transcriptomic evaluation

We evaluated the highly robust communities in an independent singe cell RNA data set of CD45+ immune cells for HCC patients from five immune-relevant sites: tumor, adjacent liver, hepatic lymph node (LN), blood, and ascites(*17*). Three patients had tumor core samples available, and we used these samples to evaluate the expression of our communities. We focused on expression in tumor core to evaluate our communities. We used Seurat to preprocess the data set(*74*). To preprocess the data, we filtered genes that were expressed in less than 40 cells and cells that had fewer than 1000, greater than 5000 genes, and cells with a proportion of transcripts mapping to mitochondrial genes greater than 5%. We then scale the data and apply PCA, then clustering and UMAP using the top 10 PCA dimensions. We use the resulting Suerat clusters and PangloaDB cell markers to identify cell marker expression(*75*). We compared the average expression of cell type markers to annotate cells and compared Seurat clusters to PangloadDB annotations to assign an immune cell type to each cluster in the core tumor samples. To evaluate the expression of our communities, and the cell-type specificity of each community, we compared the number of genes expressed from each community in each cell type to the average number of genes expressed in other cell types using a chi-squared test. We used Seurat to score the cell cycle phase of cells.

## Data Availability

All data used in the present study were openly available to the public before the initiation of the study and can be accessed online at:
https://portal.gdc.cancer.gov/
https://gtexportal.org/home/

https://portal.gdc.cancer.gov/

https://gtexportal.org/home/

## 5. Supplementary Materials

- Supplementary Note 1: SparseGMM liver communities
- Supplementary Note 2: SparseGMM model
- Supplementary Table 1: Comparison of SparseGMM to AMARETTO at different regularization values
- Supplementary Table 2: Functional enrichments for cancer, normal and shared liver modules using MSigDB collections
- Supplementary Table 3: Regulator and target genes of robust modules (average Jaccard Index=>0.7 in module subnetwork)
- Supplementary Figure 1: Co-expression patterns of target genes in highly robust communities.
- Supplementary Figure 2: Cell type identification based on Pangloa DB markers.
- Supplementary Figure 3: Comparison of gene expression in different tissues.

## Acknowledgements

We thank M. Nabian for his support on using the community AMARETTO package, and S. Napel for his advice on study design. We also thank Stanford University’s Quantitative Sciences Unit and in particular A. Gentles for their advice on statistical methods. This work supported by the National Cancer Institute (NCI) under awards: R01 CA260271, U01 CA217851 and U01 CA199241.

## Author Contributions

S.B. initiated, and designed, the study with the guidance of O.G., M.H., and K.B.; S.B. designed and developed the statistical model with the guidance from M.H. and O.G. S.B. performed all bulk and single-cell data analyses; S.B. performed all biological validation analyses; S.B. and J.A. performed entropy GSEA analysis. S.B. wrote the manuscript and generated all figures and data visualizations; All other authors reviewed and edited the manuscript.

## Supplemental Note 1

### 1. Normal liver communities

We discovered a community, which is enriched in gene sets related to the complement pathway (Supplementary Table 1). The complement system has as a critical role in immune response and liver hepatocytes are responsible for production of complement proteins. FGB, the gene encoding the beta component of fibrinogen was confirmed by LINCS perturbation data as a regulator (Table 1). Fibrinogen is a precursor of fibrin, the most abundant component of blood clots. There are many links between coagulation and innate immunity^1^, including the role of fibrin in activating the complement system^2,3^. However, this finding suggests a regulatory role of fibrin precursor in the production of complement proteins. SparseGMM also discovered CFH, a known regulator of the complement system, with no LINCS perturbation data available. CFH is a member of the regulator of complement activation gene cluster, which plays an essential role in the regulation of complement activation^4,5^.

Another highly robust community is enriched in gene sets related to digestion pathways (Supplementary Table 1). The community contains modules of the several digestive enzyme groups: lipases, liver amylase and proteases. We examined robust drivers that regulate all of the modules present in this community and used data from the LINCS perturbation library to confirms these regulatory relationships. LINCS analysis confirmed GFOD1 gene, a glucose-fructose oxidoreductase, as a regulator of the community.

### 2. Hepatocellular carcinoma communities

We discovered a community is enriched for T cell upregulated genes, showing a strong co-expression pattern of these genes in both bulk (Supplementary Figure 1) and single cell data (Figure 6). Specifically, this community is regulated by the immune checkpoint gene: PDCD1, which encodes for the PD-1 protein. The community also contains the immune checkpoint gene CTLA4, as well markers of memory CD8 resident T-Cells: CD8, CD2, CD48. CXCR3, which plays a role in T cell trafficking and function^6^, and ZNF683 a known tissue-resident lymphocyte transcription factor^7-9^ are also members of this community. Finally, this community includes T-cell receptor complex genes: CD3 epsilon, gamma, delta, and CD247 (Supplementary Table 2). Next, is a community enriched for myeloid up-regulated genes (Supplementary Figure 1, Figure 6). This community is regulated by PDCD1LG2, which encodes PD-L2, an immune checkpoint receptor ligand of PD-1. This community is also regulated by CD74, a cell surface molecule with a critical role in antigen presentation. The community also contains several members of the MHC class II genes (Supplementary Table 2).

### 3. Shared communities

First, community 21 is enriched in cell cycle gene sets. In this community, 10/30 genes were validated using LINCs perturbation data. In this shared community, cancer and normal modules shared 2 out of the 10 confirmed regulators: CDC20, a regulatory protein interacting with several other proteins at multiple points in the cell cycle^10-12^ and CDCA8 component of the chromosomal passenger complex, which is a regulator of mitosis and cell division^13-16^. The remaining 8 regulators were specific to cancer modules. We expected that most of the validated regulators would be either shared or cancer-specific, since LINCS perturbation experiments are carried out using cancer cell line data. We expect a similar regulatory pattern for the regulators discovered specifically in normal samples.

Another shared community is enriched for gene sets related to protein synthesis and formation of ribosomal subunits, ubiquitin ligase inhibitor activity and granular component. LINCS perturbation data is available for four regulators (Table 1). LINCS data analysis confirms one cancer-specific regulator, IRAK1, which does not regulate this community in normal tissue. IRAK1 plays a critical role in initiating innate immune response against foreign pathogens^17^. IRAK1 was also shown to promote cell proliferation and protect against apoptosis in HCC^18^ and to regulate the properties of liver tumor-initiating cells (TIC), including self-renewal, and tumorigenicity, suggesting IRAK1 as a potential therapeutic target of HCC^19^.

Finally, LINCS perturbation results confirmed 50% of regulators with available experimental data (3 out of 6) in the sterol biosynthesis community including ACAT2 and SREBF2 (Table 1). ACAT2, an enzyme involved in lipid metabolism and cholesterol esterification^34^, was associated with tumor growth and progression in liver^35^ and colorectal^36^ cancers. Additionally, the blockage of SREBF2, a master transcriptional regulator of cholesterol and fatty acid pathway genes was shown to completely prevent hepatocarcinogenesis^37^ and impaired the survival of HCC cell lines through the suppression of AKT-mTORC-RPS6 dependent cell proliferation^38^. This community contains 20 target genes (Supplementary Figure 1), all of which are associated with cholesterol and lipid metabolism. Thus, this community reflects both an essential metabolic liver function, as well as an accelerated sterol and lipid metabolism that is a hallmark of cancer^39^. These findings point towards an importance of sterol biosynthetic expression programs for tumor growth.

## Supplementary Note 2: Model for Sparse Gaussian Mixtures

### 1 Model

We propose a Bayesian generative model to learning the regulatory relationships among genes. In the context of gene regulatory networks, we classify genes into one of two types: target genes and regulator genes.

Regulator genes are genes undergoing genomic events that are relevant to cancer progression or tumor growth. Target genes are genes whose expression is controlled by regulator genes, and which contribute to the biological processes responsible for cancer progression. Each group of target genes is regulated by a small set of regulator genes.

This model can be formulated as follows: *X*^*T*^ = [*x*_1_*x*_2_..*x*_i_..*x*_*N*_] is a gene expression matrix *X* ∈ ℝ^*N*×*M*^, where *N* is the number of target genes and *M* is the number of subjects, *G* is a regulator expression matrix ∈ ℝ^*M*×*P*^ where *M* is the number of subjects and *P* is the number of regulator genes. Finally, *β* ∈ ℝ^*P* ×*K*^ is a weight matrix, where *K* is the number of gene modules. The mean of each Gaussian component is a vector of weights passed through a constant regulator gene expression matrix:

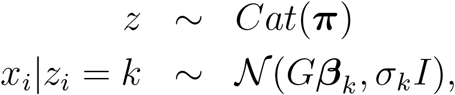

where *z*_*i*_ is the latent indicator of the mixture component that generated gene *i*. The expression of gene *i* is a sample from a Gaussian with mean equal to the weights, *β*_*k*_ passed through a constant regulator gene expression matrix *G*.*σ*_*k*_ is the variance of the Gaussian mixture component *k* and *π* is the parameter of the categorical distribution. Thus, *θ*_*k*_ = [*β*_*k*,_ *σ*_*k*_].

Our Bayesian approach combines Gaussian mixtures with 𝓁1-norm regularization to enforce sparsity on the regulator weights, resulting in a small set of regulators for each mixture component. We develop an *expectation-maximization* (EM)-based algorithm to obtain a *maximum a posteriori* (MAP) estimate the Gaussian mixture of parameters. This is detailed in the following sections.

### 2 Gaussian Mixtures for Gene Regulatory Networks

Mixture models are useful for representing data that are generated from different distributions, such as multimodal data. The data is assumed to be generated from a mixture of components, each with specific parameters that specify its distribution. The goal is to estimate these parameters using the observed data without observing the true component membership of the data points, which is a hidden or latent variable of our model. In a mixture model with *K* distributions *z*_*i*_ ∈ {1,…, *K*}, point ***x***_***i***_ is generated from distribution *k* with likelihood *p*(*x*_***i***_|*z*_*i*_ = *k*). *z*_*i*_ has the distribution *p*(*z*_*i*_) = Cat(*π*) and the K distributions are mixed as follows:

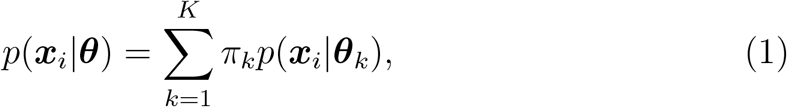

where ***θ*** are the parameters to be estimated for *k* = 1 : *K, θ* is [***θ***_**1**_ … ***θ***_***k***_ … ***θ***_***K***_]. *π*_*k*_ is the mixing weight of base distribution *k*, 0 < *π*_k_ < 1 and 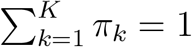. For example, a mixture of Gaussian distributions would be modeled as follows:

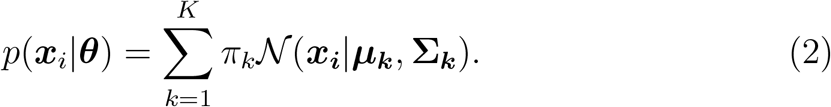

Point *i* can then be assigned to a component using the MAP or ML estimate of the parameter ***θ*** is needed.

To obtain this estimate, we fit the model for the data *𝒟*, using the iterative expectation-maximization (EM) algorithm applied to the likelihood function. The EM algorithm, consists of two steps. In the first (E) step the missing values are inferred using parameter estimates from the previous iteration. In the second (M) step, the likelihood function is maximized with respect to model parameters, giving new parameter estimates, which are improved with each subsequent iteration until convergence.

Using this model to cluster the data involves calculating the posterior probability *p*(*z*_*i*_ *= k* | *x*_***i***_, ***θ***^*t-*1^), the posterior probability that point *i* is generated from distribution *k* or the responsibility of cluster *k* for point *i*:

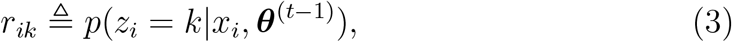

where *t* is the current iteration number. This can be expanded as:

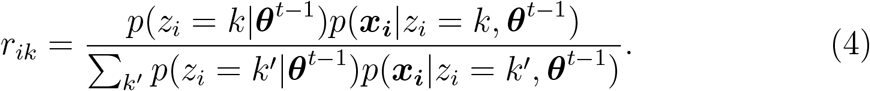

To derive the objective function, we first look at the complete log likelihood of the data, which is defined as:

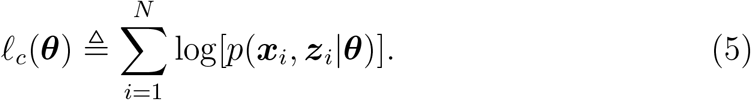

Since the cluster assignments, *z*_*i*_ are not observed the expected likelihood is used. This is defined as:

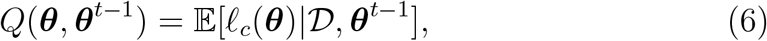

where we take the expectation to account for the fact that *z*_*i*_ is not observed.

Specifically in the case of GMM, this gives:

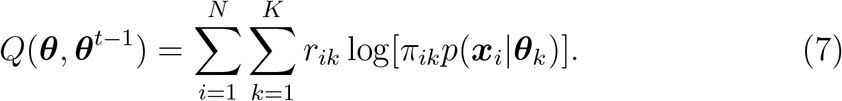

Now, a MAP estimate can be performed on the above equation in the M step, obtaining ***θ***^*t*^. In the case of Gaussian mixtures, each class conditional density is a Gaussian distribution and ***θ*** is made up of the mean and variance of each distribution and this estimate is iteratively improved. Upon convergence, the final iteration *T* gives the final estimate ***θ***^*T*^.

We can apply this model to gene regulatory networks. In this case, the average expression of target genes is the mean of the mixture component, which corresponds to a gene module. The mean of the component is a linear function of the regulator genes regulating that module. Equation (7) then becomes:

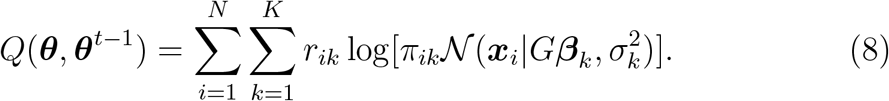

### 3 MAP Estimation - 𝓁1-norm Regularization and Sparse GMM

MAP estimation with the right prior can be useful when we would like to avoid over-fitting of parameter estimates, which can occur in the case of Maximum Likelihood Estimation (MLE). Adding parameter priors, (8) becomes:

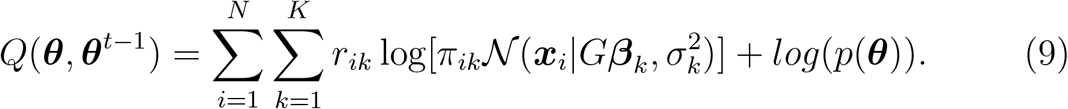

The parameters of the GMM to be estimated are ***θ***_***k***_ = [*β*_***k***,_ *σ*_***k***_] and *π*_*k*_ for *k* = 1 : *K*. In our problem, we are more interested in discovering the regulatory relationships between regulator and target genes, so we use a zero-mean Laplace prior for the weights, *β*_***k***_ and use uniform priors for *σ*_***k***_ and *π*_*k*_. Uniform priors will give the same result as MLE estimates, while the Laplace prior will give a regularized MAP estimate. Specifically, a Laplace prior is commonly chosen where a sparse solution is desired as it corresponds to *ℓ*1-norm regularization. A sparse solution can improve our understanding of gene regulatory relationships, as we hypothesize that only a few regulator genes regulate each module.

The expected likelihood function from (9) is updated to be:

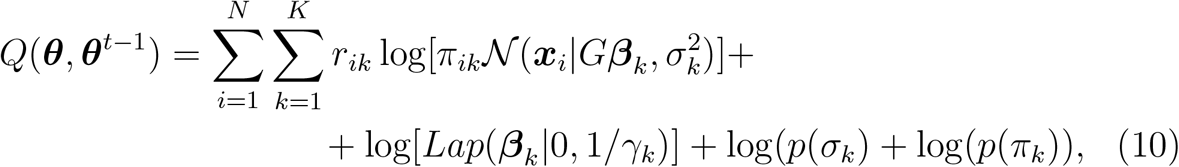

where

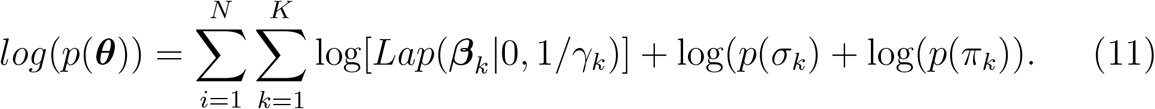

Thus, we define a sparse Gaussian Mixture model as a Gaussian mixture model, where the mean of each Gaussian component is a random vector of weights sampled from a Laplace distribution with zero mean and passed through a constant matrix.

### 4 Hierarchical Bayes modeling

Using a Laplace prior directly results in an *ℓ*1-norm, which does not give a closed form solution during optimization.

We follow an approach similar to the EM for lasso approach [1]. We then utilize the representation of a Laplace distribution as a Gaussian Scale Mixture (GSM) [2, 3].

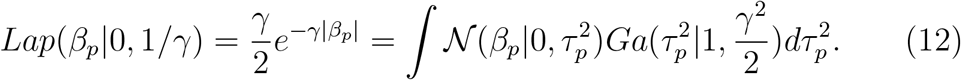

This is an example of a hierarchical Bayes model, where we include a prior on the hyperparameter *τ* ^2^ of the prior distribution *p*(***θ***). In this case, the hyperprior is the Gamma distribution with scale parameter 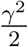. The expected complete data log likelihood is given by:

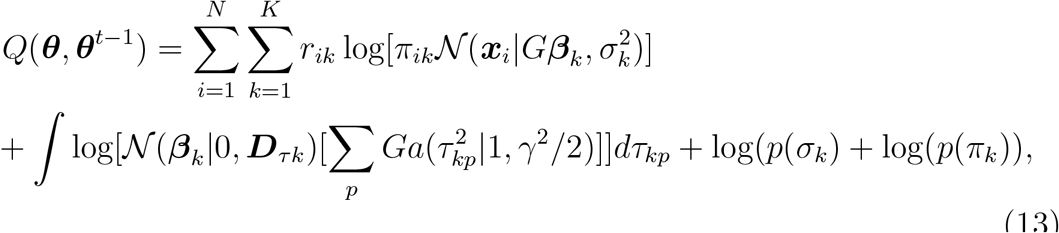

where *𝒟*_*τk*_ is *diag*(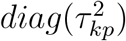) for *p* = 1 : K. The objective function then becomes:

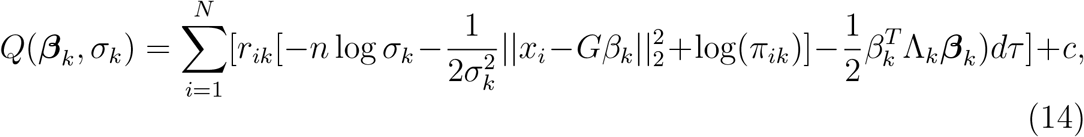

where: *D*_*ik*_ is the marginal probability of component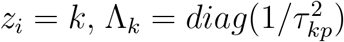 for *p* = 1 : *K* and 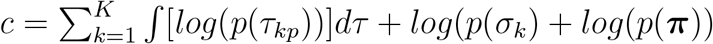.

### 5 EM Algorithm

E step: We evaluate: 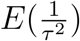 and *r*_*ik*_. From the expected complete data likelihood equation (14), the expected value of ⋀_*k*_ is

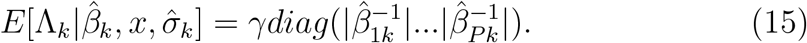

and the responsibilities are:

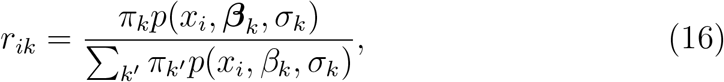

where:

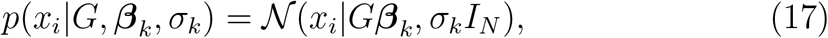

and

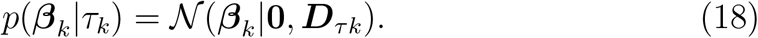

**M step:** Using a sparse learning approach [4]. We estimate model parameters *π*_*k*_, *β*_*k*_ and *σ*_*k*_ by optimizing the expected complete likelihood function with respect to each of the parameters, after substituting 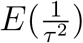 and *r*_*ik*_ obtained in the E step, taking derivative wrt *β*_*k*_

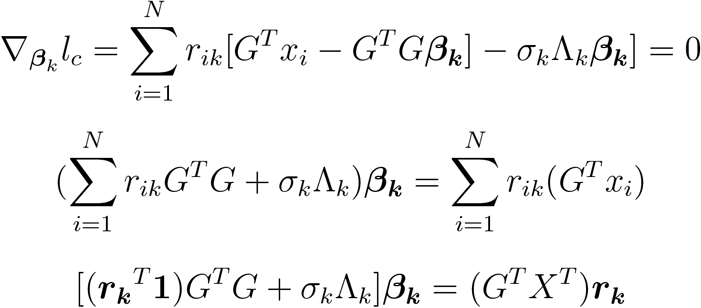

where *r*_*k*_ is the responsibility vector of component *k* ∈ *ℝ*^*N*^

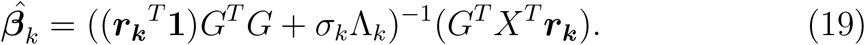

Taking the derivative wrt *σ*_*k*_ yields

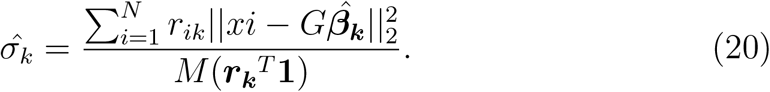

Taking the derivative wrt *π*_*k*_ yields the same result as a GMM:

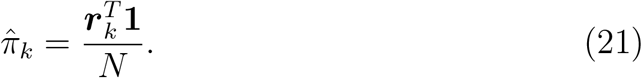

## 6 Implementation for Numerical Stability

Since we expect most *β*_*k*_ to be equal to zero, and to make the matrix under the inverse numerically stable, we use the SVD decomposition of G as follows:

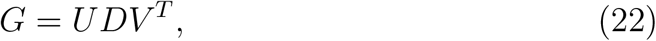

and

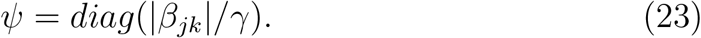

Taking the derivative wrk *β*_*k*_:

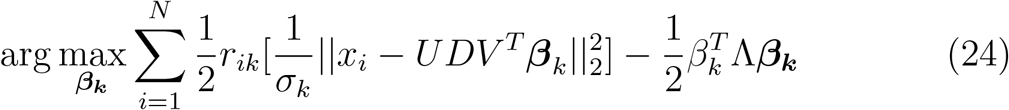

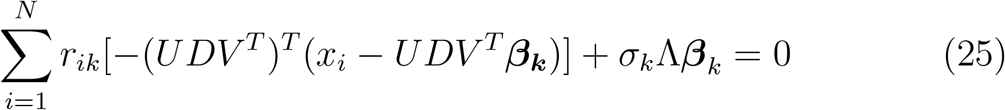

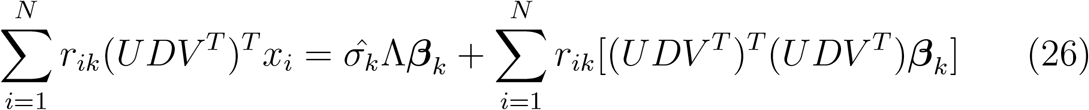

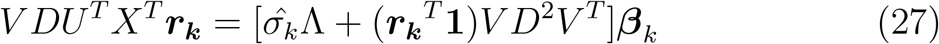

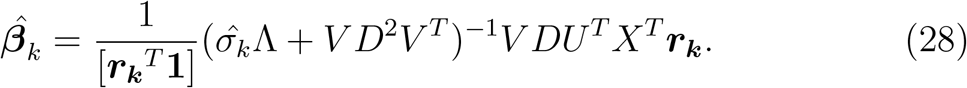

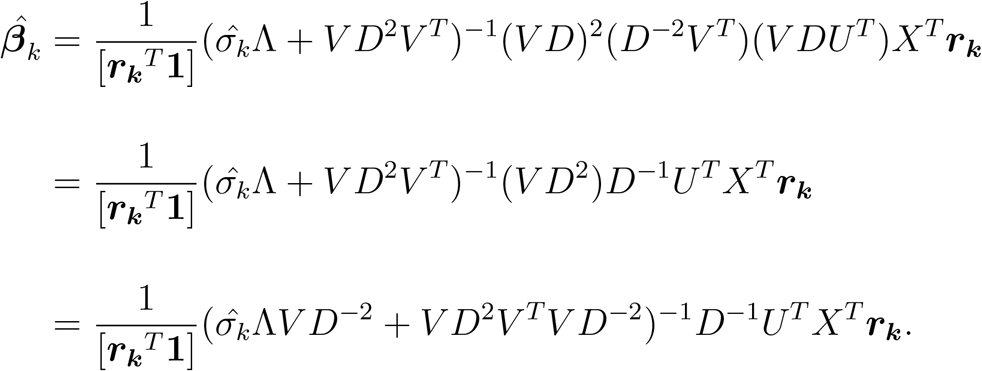

Thus, we are able to remove ⋀ from the inverse:

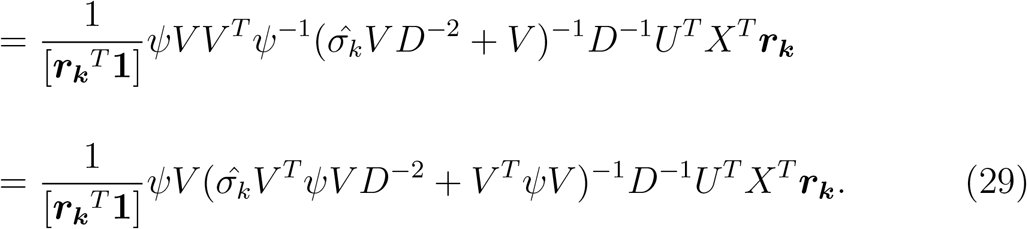

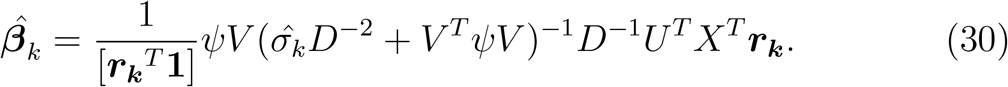

This computation of 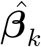 avoids numerical instability.

---

**Supplementary Figure 1:**
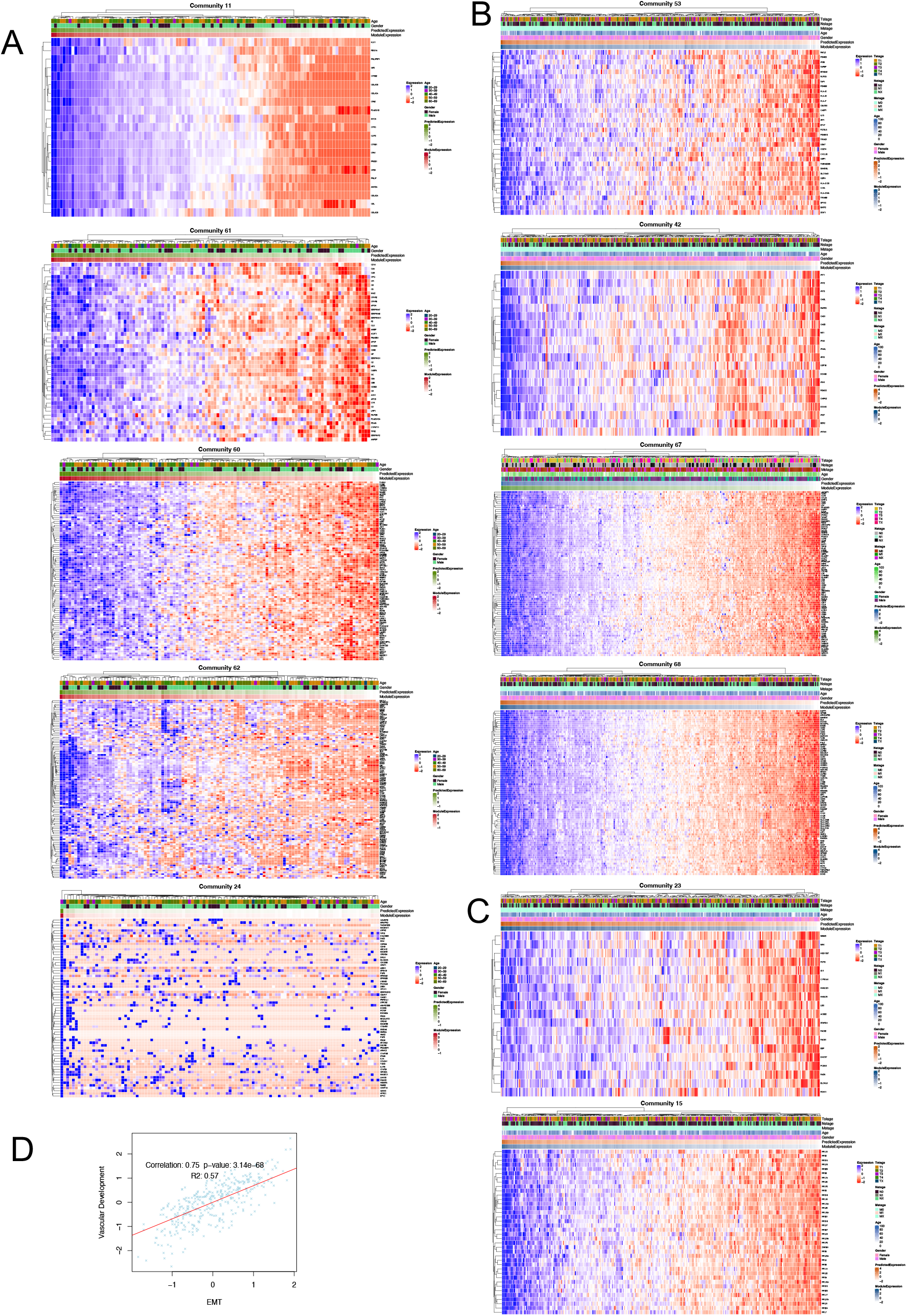
Co-expression patterns of target genes in highly robust communities. (A) normal liver communities: Lipid and protein catabolism, complement, vesicle trafficking, myofibril formation, and FGFR1 signaling. (B) Cancer communities: antigen presentation, interferon signaling, T cell and myeloid. (C) Shared communities: cell cycle and ribosome - protein synthesis. (D) Correlation between vascular development and EMT communities in TCGA samples

**Supplementary Figure 2:**
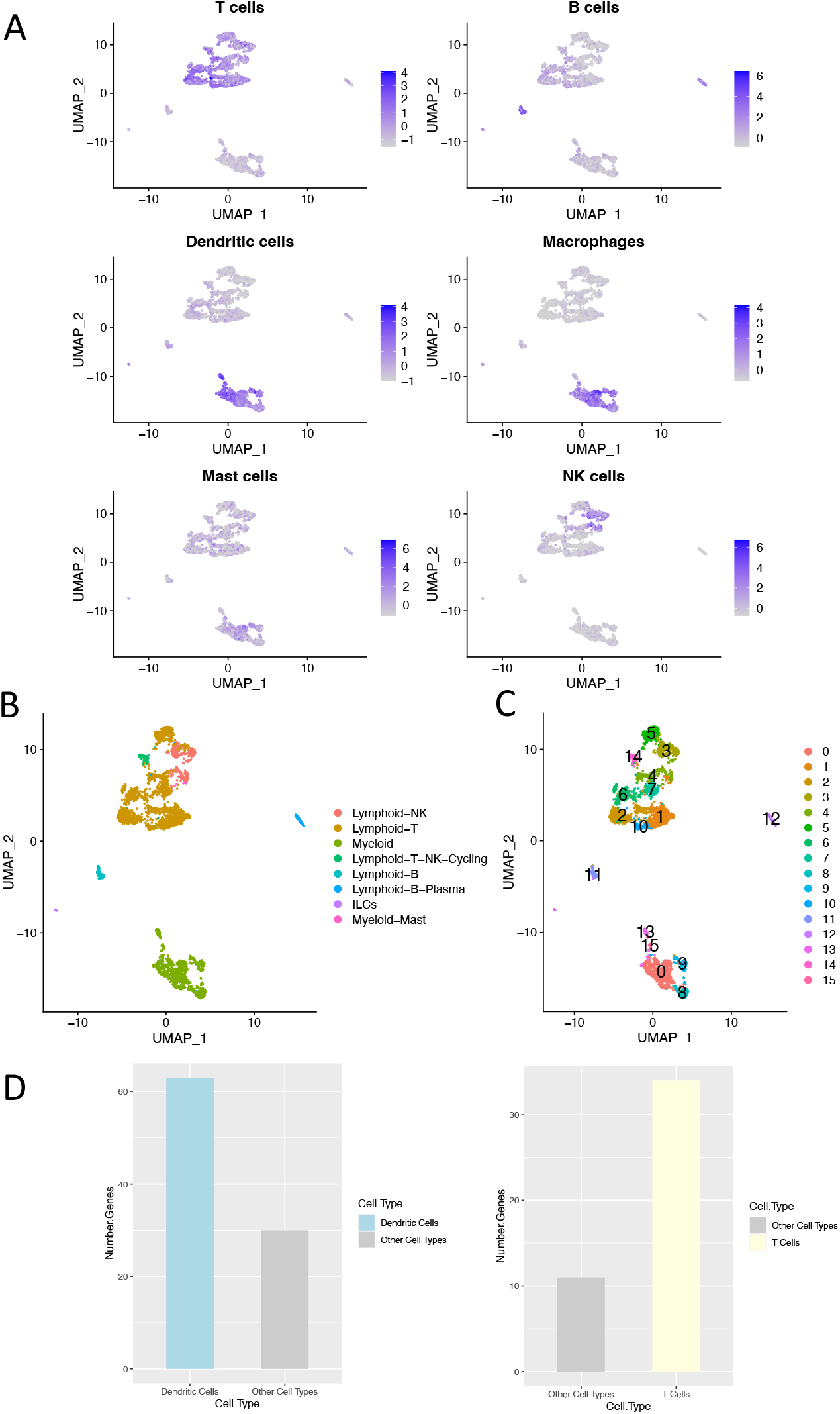
Cell type identification based on Pangloa DB markers: comparison with original data annotation and Seurat-based clustering (A) Average expression of different cell type Pangloa DB markers. (B) Original cell type assignments. (C) Seurat clusters (D) Cell type specific expression of communities 21 (dendritic cells) and 60 (T cells).

**Supplementary Figure 3:**
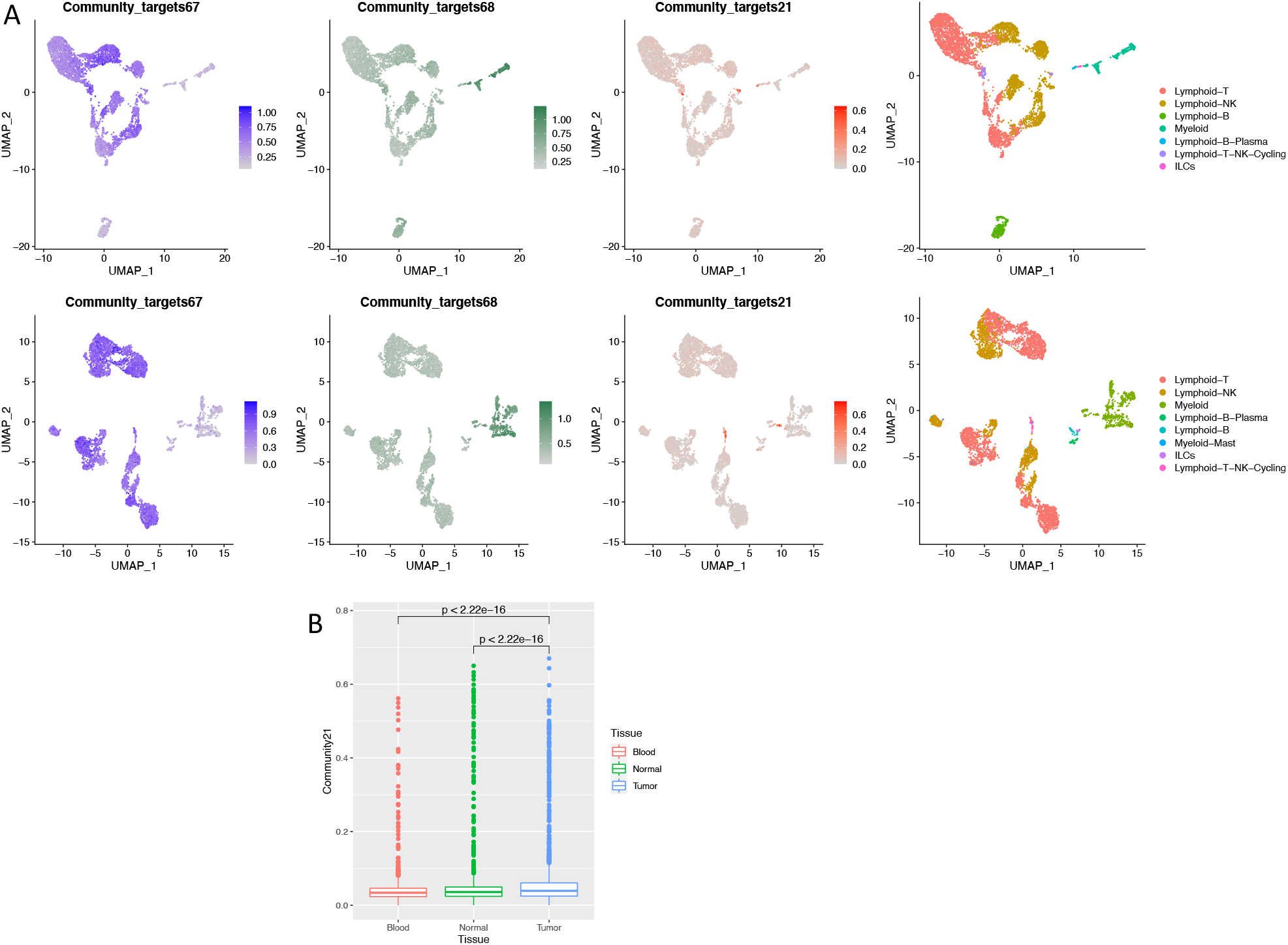
Comparison of gene expression in different tissues (A) Expression of cell-type specific robust communities. Similar expression patterns of T cell and myeloid communities occur in cancer, normal and blood tissue. Cell cycle genes are significantly more expressed in subpopulation of immune cell in tumor samples. Top, left to right expression in blood samples of target genes in T cell, myeloid and cell cycle community, and cell type assignment. Bottom, left to right normal samples of target genes in in T cell, myeloid and cell cycle community, and cell type assignment. (B) Average expression of cell cycle community target genes in blood, normal and cancer immune cells.

**Supplementary Table 1:**
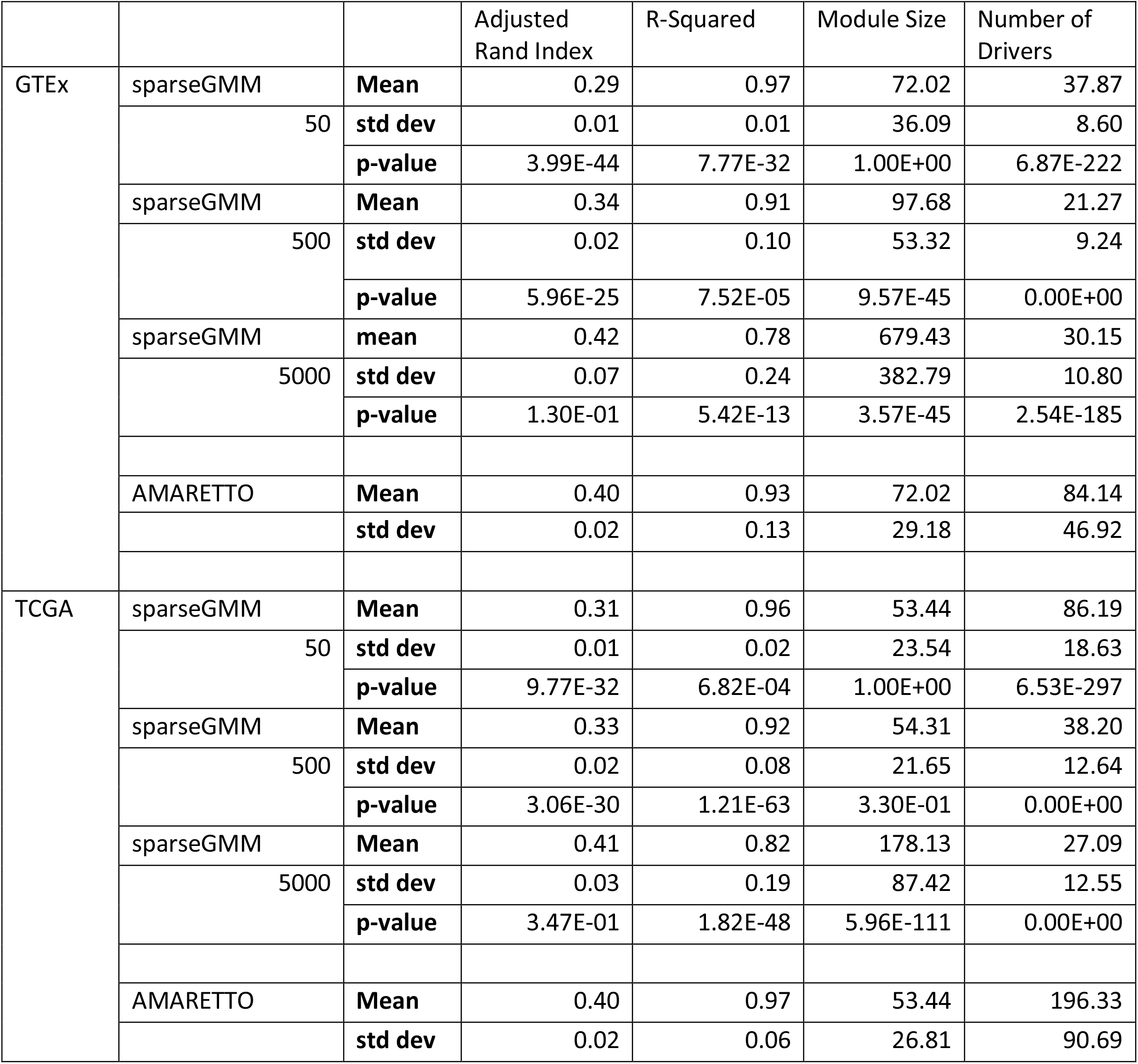
Comparison of SparseGMM to AMARETTO at different regularization values: 50, 500 and 5000. Robustness of clustering is evaluated using adjusted Rand index. Validation of regulators is represented by R-squared. Degree of sparsity is evaluated using statistics on the number of drivers. Module size informs the choice of regulatization parameter value. Standard Deviation (std dev)

## REFERENCES

1. w. b. e. Cancer Genome Atlas Research Network. Electronic address, N. Cancer Genome Atlas Research, Comprehensive and Integrative Genomic Characterization of Hepatocellular Carcinoma. Cell 169, 1327–1341 e1323 (2017).

2. G. T. Consortium, D. A. Laboratory, G. Coordinating Center-Analysis Working, G. Statistical Methods groups-Analysis Working, G. g. Enhancing, N. I. H. C. Fund, Nih/Nci, Nih/Nhgri, Nih/Nimh, Nih/Nida, N. Biospecimen Collection Source Site, R. Biospecimen Collection Source Site, V. Biospecimen Core Resource, B. Brain Bank Repository-University of Miami Brain Endowment, M. Leidos Biomedical-Project, E. Study, I. Genome Browser Data, E. B. I. Visualization, I. Genome Browser Data, U. o. C. S. C. Visualization-Ucsc Genomics Institute, a. Lead, D. A. Laboratory, C. Coordinating, N. I. H. p. management, c. Biospecimen, Pathology, Q. T. L. m. w. g. e, A. Battle, C. D. Brown, B. E. Engelhardt, S. B. Montgomery, Genetic effects on gene expression across human tissues. Nature 550, 204–213 (2017).

3. S. Xu, Y. Xu, P. Liu, S. Zhang, H. Liu, S. Slavin, S. Kumar, M. Koroleva, J. Luo, X. Wu, A. Rahman, J. Pelisek, H. Jo, S. Si, C. L. Miller, Z. G. Jin, The novel coronary artery disease risk gene JCAD/KIAA1462 promotes endothelial dysfunction and atherosclerosis. Eur Heart J 40, 2398–2408 (2019).

4. P. Mohammadi, S. E. Castel, B. B. Cummings, J. Einson, C. Sousa, P. Hoffman, S. Donkervoort, Z. Jiang, P. Mohassel, A. R. Foley, H. E. Wheeler, H. K. Im, C. G. Bonnemann, D. G. MacArthur, T. Lappalainen, Genetic regulatory variation in populations informs transcriptome analysis in rare disease. Science 366, 351–356 (2019).

5. J. D. Wang, H. S. Zhou, X. X. Tu, Y. He, Q. F. Liu, Q. Liu, Z. J. Long, Prediction of competing endogenous RNA coexpression network as prognostic markers in AML. Aging (Albany NY) 11, 3333–3347 (2019).

6. K. Basso, A. A. Margolin, G. Stolovitzky, U. Klein, R. Dalla-Favera, A. Califano, Reverse engineering of regulatory networks in human B cells. Nat Genet 37, 382–390 (2005).

7. P. Langfelder, S. Horvath, WGCNA: an R package for weighted correlation network analysis. BMC Bioinformatics 9, 559 (2008).

8. M. Hernaez, C. Blatti, O. Gevaert, Comparison of single and module-based methods for modeling gene regulatory networks. Bioinformatics 36, 558–567 (2020).

9. M. Champion, K. Brennan, T. Croonenborghs, A. J. Gentles, N. Pochet, O. Gevaert, Module Analysis Captures Pancancer Genetically and Epigenetically Deregulated Cancer Driver Genes for Smoking and Antiviral Response. EBioMedicine 27, 156–166 (2018).

10. A. Manolakos, I. Ochoa, K. Venkat, A. J. Goldsmith, O. Gevaert, CaMoDi: a new method for cancer module discovery. BMC Genomics 15 Suppl 10, S8 (2014).

11. S. Horvath, B. Zhang, M. Carlson, K. V. Lu, S. Zhu, R. M. Felciano, M. F. Laurance, W. Zhao, S. Qi, Z. Chen, Y. Lee, A. C. Scheck, L. M. Liau, H. Wu, D. H. Geschwind, P. G. Febbo, H. I. Kornblum, T. F. Cloughesy, S. F. Nelson, P. S. Mischel, Analysis of oncogenic signaling networks in glioblastoma identifies ASPM as a molecular target. Proc Natl Acad Sci U S A 103, 17402–17407 (2006).

12. U. D. Akavia, O. Litvin, J. Kim, F. Sanchez-Garcia, D. Kotliar, H. C. Causton, P. Pochanard, E. Mozes, L. A. Garraway, D. Pe’er, An integrated approach to uncover drivers of cancer. Cell 143, 1005–1017 (2010).

13. O. Gevaert, V. Villalobos, B. I. Sikic, S. K. Plevritis, Identification of ovarian cancer driver genes by using module network integration of multi-omics data. Interface Focus 3, 20130013 (2013).

14. O. Gevaert, S. Plevritis, Identifying master regulators of cancer and their downstream targets by integrating genomic and epigenomic features. Pac Symp Biocomput, 123–134 (2013).

15. A. Subramanian, P. Tamayo, V. K. Mootha, S. Mukherjee, B. L. Ebert, M. A. Gillette, A. Paulovich, S. L. Pomeroy, T. R. Golub, E. S. Lander, J. P. Mesirov, Gene set enrichment analysis: a knowledge-based approach for interpreting genome-wide expression profiles. Proc Natl Acad Sci U S A 102, 15545–15550 (2005).

16. O. Gevaert, M. Nabian, S. Bakr, C. Everaert, J. Shinde, A. Manukyan, T. Liefeld, T. Tabor, J. Xu, J. Lupberger, B. J. Haas, T. F. Baumert, M. Hernaez, M. Reich, F. J. Quintana, E. J. Uhlmann, A. M. Krichevsky, J. P. Mesirov, V. Carey, N. Pochet, Imaging-AMARETTO: An Imaging Genomics Software Tool to Interrogate Multiomics Networks for Relevance to Radiography and Histopathology Imaging Biomarkers of Clinical Outcomes. JCO Clin Cancer Inform 4, 421–435 (2020).

17. Q. Zhang, Y. He, N. Luo, S. J. Patel, Y. Han, R. Gao, M. Modak, S. Carotta, C. Haslinger, D. Kind, G. W. Peet, G. Zhong, S. Lu, W. Zhu, Y. Mao, M. Xiao, M. Bergmann, X. Hu, S. P. Kerkar, A. B. Vogt, S. Pflanz, K. Liu, J. Peng, X. Ren, Z. Zhang, Landscape and Dynamics of Single Immune Cells in Hepatocellular Carcinoma. Cell 179, 829–845 e820 (2019).

18. R. D. Leclerc, Survival of the sparsest: robust gene networks are parsimonious. Mol Syst Biol 4, 213 (2008).

19. I. Sansal, E. Dupont, D. Toru, C. Evrard, P. Rouget, NPDC-1, a regulator of neural cell proliferation and differentiation, interacts with E2F-1, reduces its binding to DNA and modulates its transcriptional activity. Oncogene 19, 5000–5009 (2000).

20. L. V. Mohan Rao, C. T. Esmon, U. R. Pendurthi, Endothelial cell protein C receptor: a multiliganded and multifunctional receptor. Blood 124, 1553–1562 (2014).

21. M. Bezuhly, R. Cullen, C. T. Esmon, S. F. Morris, K. A. West, B. Johnston, R. S. Liwski, Role of activated protein C and its receptor in inhibition of tumor metastasis. Blood 113, 3371–3374 (2009).

22. G. L. Van Sluis, T. M. Niers, C. T. Esmon, W. Tigchelaar, D. J. Richel, H. R. Buller, C. J. Van Noorden, C. A. Spek, Endogenous activated protein C limits cancer cell extravasation through sphingosine-1-phosphate receptor 1-mediated vascular endothelial barrier enhancement. Blood 114, 1968–1973 (2009).

23. L. Yang, S. Yue, L. Yang, X. Liu, Z. Han, Y. Zhang, L. Li, Sphingosine kinase/sphingosine 1-phosphate (S1P)/S1P receptor axis is involved in liver fibrosis-associated angiogenesis. J Hepatol 59, 114–123 (2013).

24. A. Ben Shoham, G. Malkinson, S. Krief, Y. Shwartz, Y. Ely, N. Ferrara, K. Yaniv, E. Zelzer, S1P1 inhibits sprouting angiogenesis during vascular development. Development 139, 3859–3869 (2012).

25. V. A. Balaji Ragunathrao, M. Anwar, M. Z. Akhter, A. Chavez, Y. Mao, V. Natarajan, S. Lakshmikanthan, M. Chrzanowska-Wodnicka, A. Z. Dudek, L. Claesson-Welsh, J. K. Kitajewski, K. K. Wary, A. B. Malik, D. Mehta, Sphingosine-1-Phosphate Receptor 1 Activity Promotes Tumor Growth by Amplifying VEGF-VEGFR2 Angiogenic Signaling. Cell Rep 29, 3472–3487 e3474 (2019).

26. A. Cartier, T. Leigh, C. H. Liu, T. Hla, Endothelial sphingosine 1-phosphate receptors promote vascular normalization and antitumor therapy. Proc Natl Acad Sci U S A 117, 3157–3166 (2020).

27. M. Uchiba, K. Okajima, Y. Oike, Y. Ito, K. Fukudome, H. Isobe, T. Suda, Activated protein C induces endothelial cell proliferation by mitogen-activated protein kinase activation in vitro and angiogenesis in vivo. Circ Res 95, 34–41 (2004).

28. Q. C. Yu, W. Song, D. Wang, Y. A. Zeng, Identification of blood vascular endothelial stem cells by the expression of protein C receptor. Cell Res 26, 1079–1098 (2016).

29. E. Ducros, S. Mirshahi, D. Azzazene, S. Camilleri-Broet, E. Mery, H. Al Farsi, H. Althawadi, S. Besbess, J. Chidiac, E. Pujade-Lauraine, A. Therwath, J. Soria, M. Mirshahi, Endothelial protein C receptor expressed by ovarian cancer cells as a possible biomarker of cancer onset. Int J Oncol 41, 433–440 (2012).

30. H. J. Choi, S. S. Rho, D. H. Choi, Y. G. Kwon, LDB2 regulates the expression of DLL4 through the formation of oligomeric complexes in endothelial cells. BMB Rep 51, 21–26 (2018).

31. R. Brutsch, S. S. Liebler, J. Wustehube, A. Bartol, S. E. Herberich, M. G. Adam, A. Telzerow, H. G. Augustin, A. Fischer, Integrin cytoplasmic domain-associated protein-1 attenuates sprouting angiogenesis. Circ Res 107, 592–601 (2010).

32. Y. Nemerson, The role of lipids in the tissue factor pathway of blood coagulation. Adv Exp Med Biol 63, 245–253 (1975).

33. A. J. Marcus, The role of lipids in blood coagulation. Adv Lipid Res 4, 1–37 (1966).

34. Y. Inoue, L. L. Peters, S. H. Yim, J. Inoue, F. J. Gonzalez, Role of hepatocyte nuclear factor 4alpha in control of blood coagulation factor gene expression. J Mol Med (Berl) 84, 334–344 (2006).

35. H. Safdar, K. L. Cheung, H. L. Vos, F. J. Gonzalez, P. H. Reitsma, Y. Inoue, B. J. van Vlijmen, Modulation of mouse coagulation gene transcription following acute in vivo delivery of synthetic small interfering RNAs targeting HNF4alpha and C/EBPalpha. PLoS One 7, e38104 (2012).

36. H. Safdar, Y. Inoue, G. H. van Puijvelde, P. H. Reitsma, B. J. van Vlijmen, The role of hepatocyte nuclear factor 4alpha in regulating mouse hepatic anticoagulation and fibrinolysis gene transcript levels. J Thromb Haemost 8, 2839–2841 (2010).

37. A. DeLaForest, F. Di Furio, R. Jing, A. Ludwig-Kubinski, K. Twaroski, A. Urick, K. Pulakanti, S. Rao, S. A. Duncan, HNF4A Regulates the Formation of Hepatic Progenitor Cells from Human iPSC-Derived Endoderm by Facilitating Efficient Recruitment of RNA Pol II. Genes (Basel) 10, (2018).

38. M. Qu, T. Duffy, T. Hirota, S. A. Kay, Nuclear receptor HNF4A transrepresses CLOCK:BMAL1 and modulates tissue-specific circadian networks. Proc Natl Acad Sci U S A 115, E12305–E12312 (2018).

39. H. Nakajima, T. Tanoue, Lulu2 regulates the circumferential actomyosin tensile system in epithelial cells through p114RhoGEF. J Cell Biol 195, 245–261 (2011).

40. D. C. Bosanquet, L. Ye, K. G. Harding, W. G. Jiang, Expressed in high metastatic cells (Ehm2) is a positive regulator of keratinocyte adhesion and motility: The implication for wound healing. J Dermatol Sci 71, 115–121 (2013).

41. R. Szabo, S. Netzel-Arnett, J. P. Hobson, T. M. Antalis, T. H. Bugge, Matriptase-3 is a novel phylogenetically preserved membrane-anchored serine protease with broad serpin reactivity. Biochem J 390, 231–242 (2005).

42. H. Rubin, Serine protease inhibitors (SERPINS): where mechanism meets medicine. Nat Med 2, 632–633 (1996).

43. P. Adami, T. M. Duncan, J. O. McIntyre, C. E. Carter, C. Fu, M. Melin, N. Latruffe, S. Fleischer, Monoclonal antibodies for structure-function studies of (R)-3-hydroxybutyrate dehydrogenase, a lipid-dependent membrane-bound enzyme. Biochem J 292 (Pt 3), 863–872 (1993).

44. M. J. Bennett, L. K. Russell, C. Tokunaga, S. B. Narayan, L. Tan, A. Seegmiller, R. L. Boriack, A. W. Strauss, Reye-like syndrome resulting from novel missense mutations in mitochondrial medium-and short-chain l-3-hydroxy-acyl-CoA dehydrogenase. Mol Genet Metab 89, 74–79 (2006).

45. P. T. Clayton, S. Eaton, A. Aynsley-Green, M. Edginton, K. Hussain, S. Krywawych, V. Datta, H. E. Malingre, R. Berger, I. E. van den Berg, Hyperinsulinism in short-chain L-3-hydroxyacyl-CoA dehydrogenase deficiency reveals the importance of beta-oxidation in insulin secretion. J Clin Invest 108, 457–465 (2001).

46. J. J. Barycki, L. K. O’Brien, J. M. Bratt, R. Zhang, R. Sanishvili, A. W. Strauss, L. J. Banaszak, Biochemical characterization and crystal structure determination of human heart short chain L-3-hydroxyacyl-CoA dehydrogenase provide insights into catalytic mechanism. Biochemistry 38, 5786–5798 (1999).

47. G. A. Webster, N. D. Perkins, Transcriptional cross talk between NF-kappaB and p53. Mol Cell Biol 19, 3485–3495 (1999).

48. G. Schneider, A. Henrich, G. Greiner, V. Wolf, A. Lovas, M. Wieczorek, T. Wagner, S. Reichardt, A. von Werder, R. M. Schmid, F. Weih, T. Heinzel, D. Saur, O. H. Kramer, Cross talk between stimulated NF-kappaB and the tumor suppressor p53. Oncogene 29, 2795–2806 (2010).

49. C. Berger, Y. Qian, X. Chen, The p53-estrogen receptor loop in cancer. Curr Mol Med 13, 1229–1240 (2013).

50. J. M. A. Delou, A. S. O. Souza, L. C. M. Souza, H. L. Borges, Highlights in Resistance Mechanism Pathways for Combination Therapy. Cells 8, (2019).

51. L. M. Ellis, D. J. Hicklin, Resistance to Targeted Therapies: Refining Anticancer Therapy in the Era of Molecular Oncology. Clin Cancer Res 15, 7471–7478 (2009).

52. K. Schulze, S. Imbeaud, E. Letouze, L. B. Alexandrov, J. Calderaro, S. Rebouissou, G. Couchy, C. Meiller, J. Shinde, F. Soysouvanh, A. L. Calatayud, R. Pinyol, L. Pelletier, C. Balabaud, A. Laurent, J. F. Blanc, V. Mazzaferro, F. Calvo, A. Villanueva, J. C. Nault, P. Bioulac-Sage, M. R. Stratton, J. M. Llovet, J. Zucman-Rossi, Exome sequencing of hepatocellular carcinomas identifies new mutational signatures and potential therapeutic targets. Nat Genet 47, 505–511 (2015).

53. J. D. Yang, P. Hainaut, G. J. Gores, A. Amadou, A. Plymoth, L. R. Roberts, A global view of hepatocellular carcinoma: trends, risk, prevention and management. Nat Rev Gastroenterol Hepatol 16, 589–604 (2019).

54. C. Ma, M. Han, B. Heinrich, Q. Fu, Q. Zhang, M. Sandhu, D. Agdashian, M. Terabe, J. A. Berzofsky, V. Fako, T. Ritz, T. Longerich, C. M. Theriot, J. A. McCulloch, S. Roy, W. Yuan, V. Thovarai, S. K. Sen, M. Ruchirawat, F. Korangy, X. W. Wang, G. Trinchieri, T. F. Greten, Gut microbiome-mediated bile acid metabolism regulates liver cancer via NKT cells. Science 360, (2018).

55. M. Kalra, J. Mayes, S. Assefa, A. K. Kaul, R. Kaul, Role of sex steroid receptors in pathobiology of hepatocellular carcinoma. World J Gastroenterol 14, 5945–5961 (2008).

56. M. G. Ghosh, D. A. Thompson, R. J. Weigel, PDZK1 and GREB1 are estrogen-regulated genes expressed in hormone-responsive breast cancer. Cancer Res 60, 6367–6375 (2000).

57. K. Shostak, F. Patrascu, S. I. Goktuna, P. Close, L. Borgs, L. Nguyen, F. Olivier, A. Rammal, H. Brinkhaus, M. Bentires-Alj, J. C. Marine, A. Chariot, MDM2 restrains estrogen-mediated AKT activation by promoting TBK1-dependent HPIP degradation. Cell Death Differ 21, 811–824 (2014).

58. R. C. Baxter, IGF binding proteins in cancer: mechanistic and clinical insights. Nat Rev Cancer 14, 329–341 (2014).

59. A. Grimberg, C. M. Coleman, T. F. Burns, B. P. Himelstein, C. J. Koch, P. Cohen, W. S. El-Deiry, p53-Dependent and p53-independent induction of insulin-like growth factor binding protein-3 by deoxyribonucleic acid damage and hypoxia. J Clin Endocrinol Metab 90, 3568–3574 (2005).

60. O. Neumann, M. Kesselmeier, R. Geffers, R. Pellegrino, B. Radlwimmer, K. Hoffmann, V. Ehemann, P. Schemmer, P. Schirmacher, J. Lorenzo Bermejo, T. Longerich, Methylome analysis and integrative profiling of human HCCs identify novel protumorigenic factors. Hepatology 56, 1817–1827 (2012).

61. Z. M. Shao, M. S. Sheikh, J. V. Ordonez, P. Feng, T. Kute, J. C. Chen, S. Aisner, L. Schnaper, D. LeRoith, C. T. Roberts, Jr., et al., IGFBP-3 gene expression and estrogen receptor status in human breast carcinoma. Cancer Res 52, 5100–5103 (1992).

62. R. L. Rocha, S. G. Hilsenbeck, J. G. Jackson, A. V. Lee, J. A. Figueroa, D. Yee, Correlation of insulin-like growth factor-binding protein-3 messenger RNA with protein expression in primary breast cancer tissues: detection of higher levels in tumors with poor prognostic features. J Natl Cancer Inst 88, 601–606 (1996).

63. J. A. Figueroa, J. G. Jackson, W. L. McGuire, R. F. Krywicki, D. Yee, Expression of insulin-like growth factor binding proteins in human breast cancer correlates with estrogen receptor status. J Cell Biochem 52, 196–205 (1993).

64. H. Yu, M. A. Levesque, M. J. Khosravi, A. Papanastasiou-Diamandi, G. M. Clark, E. P. Diamandis, Associations between insulin-like growth factors and their binding proteins and other prognostic indicators in breast cancer. Br J Cancer 74, 1242–1247 (1996).

65. D. Nonaka, L. Chiriboga, R. A. Soslow, Expression of pax8 as a useful marker in distinguishing ovarian carcinomas from mammary carcinomas. Am J Surg Pathol 32, 1566–1571 (2008).

66. M. Ohmori, N. Harii, T. Endo, T. Onaya, Tumor necrosis factor-alpha regulation of thyroid transcription factor-1 and Pax-8 in rat thyroid FRTL-5 cells. Endocrinology 140, 4651–4658 (1999).

67. D. Ghannam-Shahbari, E. Jacob, R. R. Kakun, T. Wasserman, L. Korsensky, O. Sternfeld, J. Kagan, D. R. Bublik, S. Aviel-Ronen, K. Levanon, E. Sabo, S. Larisch, M. Oren, D. Hershkovitz, R. Perets, PAX8 activates a p53-p21-dependent pro-proliferative effect in high grade serous ovarian carcinoma. Oncogene 37, 2213–2224 (2018).

68. J. M. Lowe, D. Menendez, P. R. Bushel, M. Shatz, E. L. Kirk, M. A. Troester, S. Garantziotis, M. B. Fessler, M. A. Resnick, p53 and NF-kappaB coregulate proinflammatory gene responses in human macrophages. Cancer Res 74, 2182–2192 (2014).

69. G. Di Minin, A. Bellazzo, M. Dal Ferro, G. Chiaruttini, S. Nuzzo, S. Bicciato, S. Piazza, D. Rami, R. Bulla, R. Sommaggio, A. Rosato, G. Del Sal, L. Collavin, Mutant p53 reprograms TNF signaling in cancer cells through interaction with the tumor suppressor DAB2IP. Mol Cell 56, 617–629 (2014).

70. Q. Wang, J. Armenia, C. Zhang, A. V. Penson, E. Reznik, L. Zhang, T. Minet, A. Ochoa, B. E. Gross, C. A. Iacobuzio-Donahue, D. Betel, B. S. Taylor, J. Gao, N. Schultz, Unifying cancer and normal RNA sequencing data from different sources. Sci Data 5, 180061 (2018).

71. B. J. Stubbs, S. Gopaulakrishnan, K. Glass, N. Pochet, C. Everaert, B. Raby, V. Carey, TFutils: Data structures for transcription factor bioinformatics. F1000Res 8, 152 (2019).

72. C. H. Mermel, S. E. Schumacher, B. Hill, M. L. Meyerson, R. Beroukhim, G. Getz, GISTIC2.0 facilitates sensitive and confident localization of the targets of focal somatic copy-number alteration in human cancers. Genome Biol 12, R41 (2011).

73. O. Gevaert, MethylMix: an R package for identifying DNA methylation-driven genes. Bioinformatics 31, 1839–1841 (2015).

74. T. Stuart, A. Butler, P. Hoffman, C. Hafemeister, E. Papalexi, W. M. Mauck, 3rd, Y. Hao, M. Stoeckius, P. Smibert, R. Satija, Comprehensive Integration of Single-Cell Data. Cell 177, 1888–1902 e1821 (2019).

75. O. Franzen, L. M. Gan, J. L. M. Bjorkegren, PanglaoDB: a web server for exploration of mouse and human single-cell RNA sequencing data. Database (Oxford) 2019, (2019).

## References

1 Pahlman, L. I. et al. Antimicrobial activity of fibrinogen and fibrinogen-derived peptides--a novel link between coagulation and innate immunity. Thromb Haemost 109, 930–939, doi:10.1160/TH12-10-0739 (2013).

2 Endo, Y. et al. Interactions of ficolin and mannose-binding lectin with fibrinogen/fibrin augment the lectin complement pathway. J Innate Immun 2, 33–42, doi:10.1159/000227805 (2010).

3 Hoppe, B. Fibrinogen and factor XIII at the intersection of coagulation, fibrinolysis and inflammation. Thromb Haemost 112, 649–658, doi:10.1160/TH14-01-0085 (2014).

4 Kajander, T. et al. Dual interaction of factor H with C3d and glycosaminoglycans in host-nonhost discrimination by complement. Proc Natl Acad Sci U S A 108, 2897–2902, doi:10.1073/pnas.1017087108 (2011).

5 Blaum, B. S. et al. Structural basis for sialic acid-mediated self-recognition by complement factor H. Nat Chem Biol 11, 77–82, doi:10.1038/nchembio.1696 (2015).

6 Groom, J. R. & Luster, A. D. CXCR3 in T cell function. Exp Cell Res 317, 620–631, doi:10.1016/j.yexcr.2010.12.017 (2011).

7 van Gisbergen, K. P. et al. Mouse Hobit is a homolog of the transcriptional repressor Blimp-1 that regulates NKT cell effector differentiation. Nat Immunol 13, 864–871, doi:10.1038/ni.2393 (2012).

8 Mackay, L. K. et al. Hobit and Blimp1 instruct a universal transcriptional program of tissue residency in lymphocytes. Science 352, 459–463, doi:10.1126/science.aad2035 (2016).

9 Bird, L. Lymphocyte responses: Hunker down with HOBIT and BLIMP1. Nat Rev Immunol 16, 338–339, doi:10.1038/nri.2016.61 (2016).

10 Fang, G., Yu, H. & Kirschner, M. W. Direct binding of CDC20 protein family members activates the anaphase-promoting complex in mitosis and G1. Mol Cell 2, 163–171, doi:10.1016/s1097-2765(00)80126-4 (1998).

11 Fang, G., Yu, H. & Kirschner, M. W. The checkpoint protein MAD2 and the mitotic regulator CDC20 form a ternary complex with the anaphase-promoting complex to control anaphase initiation. Genes Dev 12, 1871–1883, doi:10.1101/gad.12.12.1871 (1998).

12 Kramer, E. R., Gieffers, C., Holzl, G., Hengstschlager, M. & Peters, J. M. Activation of the human anaphase-promoting complex by proteins of the CDC20/Fizzy family. Curr Biol 8, 1207–1210, doi:10.1016/s0960-9822(07)00510-6 (1998).

13 Jelluma, N. et al. Mps1 phosphorylates Borealin to control Aurora B activity and chromosome alignment. Cell 132, 233–246, doi:10.1016/j.cell.2007.11.046 (2008).

14 Klein, U. R., Nigg, E. A. & Gruneberg, U. Centromere targeting of the chromosomal passenger complex requires a ternary subcomplex of Borealin, Survivin, and the N-terminal domain of INCENP. Mol Biol Cell 17, 2547–2558, doi:10.1091/mbc.e05-12-1133 (2006).

15 Gassmann, R. et al. Borealin: a novel chromosomal passenger required for stability of the bipolar mitotic spindle. J Cell Biol 166, 179–191, doi:10.1083/jcb.200404001 (2004).

16 Sampath, S. C. et al. The chromosomal passenger complex is required for chromatin-induced microtubule stabilization and spindle assembly. Cell 118, 187–202, doi:10.1016/j.cell.2004.06.026 (2004).

17 Wesche, H. et al. IRAK-M is a novel member of the Pelle/interleukin-1 receptor-associated kinase (IRAK) family. J Biol Chem 274, 19403–19410, doi:10.1074/jbc.274.27.19403 (1999).

18 Li, N. et al. Targeting interleukin-1 receptor-associated kinase 1 for human hepatocellular carcinoma. J Exp Clin Cancer Res 35, 140, doi:10.1186/s13046-016-0413-0 (2016).

19 Cheng, B. Y. et al. IRAK1 Augments Cancer Stemness and Drug Resistance via the AP-1/AKR1B10 Signaling Cascade in Hepatocellular Carcinoma. Cancer Res 78, 2332–2342, doi:10.1158/0008-5472.CAN-17-2445 (2018).

20 Sansal, I., Dupont, E., Toru, D., Evrard, C. & Rouget, P. NPDC-1, a regulator of neural cell proliferation and differentiation, interacts with E2F-1, reduces its binding to DNA and modulates its transcriptional activity. Oncogene 19, 5000–5009, doi:10.1038/sj.onc.1203843 (2000).

21 Bezuhly, M. et al. Role of activated protein C and its receptor in inhibition of tumor metastasis. Blood 113, 3371–3374, doi:10.1182/blood-2008-05-159434 (2009).

22 Van Sluis, G. L. et al. Endogenous activated protein C limits cancer cell extravasation through sphingosine-1-phosphate receptor 1-mediated vascular endothelial barrier enhancement. Blood 114, 1968–1973, doi:10.1182/blood-2009-04-217679 (2009).

23 Yang, L. et al. Sphingosine kinase/sphingosine 1-phosphate (S1P)/S1P receptor axis is involved in liver fibrosis-associated angiogenesis. J Hepatol 59, 114–123, doi:10.1016/j.jhep.2013.02.021 (2013).

24 Ben Shoham, A. et al. S1P1 inhibits sprouting angiogenesis during vascular development. Development 139, 3859–3869, doi:10.1242/dev.078550 (2012).

25 Balaji Ragunathrao, V. A. et al. Sphingosine-1-Phosphate Receptor 1 Activity Promotes Tumor Growth by Amplifying VEGF-VEGFR2 Angiogenic Signaling. Cell Rep 29, 3472–3487 e3474, doi:10.1016/j.celrep.2019.11.036 (2019).

26 Cartier, A., Leigh, T., Liu, C. H. & Hla, T. Endothelial sphingosine 1-phosphate receptors promote vascular normalization and antitumor therapy. Proc Natl Acad Sci U S A 117, 3157–3166, doi:10.1073/pnas.1906246117 (2020).

27 Choi, H. J., Rho, S. S., Choi, D. H. & Kwon, Y. G. LDB2 regulates the expression of DLL4 through the formation of oligomeric complexes in endothelial cells. BMB Rep 51, 21–26, doi:10.5483/bmbrep.2018.51.1.140 (2018).

28 Brutsch, R. et al. Integrin cytoplasmic domain-associated protein-1 attenuates sprouting angiogenesis. Circ Res 107, 592–601, doi:10.1161/CIRCRESAHA.110.217257 (2010).

29 Cancer Genome Atlas Research Network. Electronic address, w. b. e. & Cancer Genome Atlas Research, N. Comprehensive and Integrative Genomic Characterization of Hepatocellular Carcinoma. Cell 169, 1327–1341 e1323, doi:10.1016/j.cell.2017.05.046 (2017).

## References

[1] K. P. Murphy, Machine learning: a probabilistic perspective. MIT press, 2012.

[2] D. F. Andrews and C. L. Mallows, “Scale mixtures of normal distributions,” Journal of the Royal Statistical Society: Series B (Methodological), vol. 36, no. 1, pp. 99–102, 1974.

[3] M. West, “On scale mixtures of normal distributions,” Biometrika, vol. 74, no. 3, pp. 646–648, 1987.

[4] M. A. Figueiredo, “Adaptive sparseness for supervised learning,” IEEE transactions on pattern analysis and machine intelligence, vol. 25, no. 9, pp. 1150–1159, 2003.

